# Availability and Use of Mobile Health Technology for Disease Diagnosis and Treatment Support by Health Workers in the Ashanti Region of Ghana: A Cross-sectional Survey

**DOI:** 10.1101/2021.05.04.21256622

**Authors:** Ernest Osei, Kwasi Agyei, Boikhutso Tlou, Tivani P. Mashamba-Thompson

## Abstract

**Background:** Mobile health (mHealth) technologies have been identified as promising strategies for improving access to healthcare delivery and patient outcomes. However, the extent of availability and use of mHealth among healthcare professionals in Ghana is not known. The main objective of this study is to determine the availability and use of mHealth for disease diagnosis and treatment support by health professionals in the Ashanti Region of Ghana.

**Methods:** A cross-sectional survey was carried out among 285 healthcare professionals across 100 primary healthcare clinics in the Ashanti Region, Ghana. We obtained data on the participants’ background, available health infrastructure, healthcare workforce competency, ownership of a mobile wireless device, usefulness of mHealth, ease of use of mHealth, user satisfaction, and behavioural intention to use mHealth. Descriptive statistics were conducted to characterize healthcare professionals’ demographics and clinical features. Multivariate logistic regression analysis was performed to explore the influence of the demographic factors on the availability and use of mHealth for disease diagnosis and treatment support. STATA version 15 was used to compute all the statistical analyses.

**Findings:** Out of the 285 healthcare professionals, 62.8% indicated that mHealth applications are available to them, while 37.2% had no access to mHealth. Of the 185 healthcare professionals who had access to mHealth, 98.4% are currently using mHealth to support healthcare delivery. Logistic regression model analysis significantly (p< 0.05) identified factors associated with the availability and use of mHealth applications for disease diagnosis and treatment support. There was a significant association between the availability and use of mHealth for disease diagnosis and treatment support from the chi-square test analysis.

**Conclusion:** The findings demonstrate a low-level use of mHealth for disease diagnosis and treatment support by healthcare professionals at the rural primary healthcare clinics. We encourage policymakers to promote the implementation of mHealth in rural primary health clinics.

**Key questions:** *What is already known:*

- Digitizing healthcare systems with mobile health technologies have been identified as essential tools for improving access to healthcare delivery in sub-Saharan Africa.
- In Ghana, mobile phones and their applications’ availability and utilization as of 2018 was estimated to be about 52% and is expected to increase steadily.
- Ghana has given considerable attention to mobile health technologies and applications’ role in transforming healthcare delivery.

*What are the new findings:*

- The study reveals that 63% of healthcare professionals indicated that mHealth applications are available to them, while 37% do not have access to mHealth applications.
- The study results illustrate that healthcare professionals primarily use mHealth applications to screen or diagnose existing many disease conditions in Ghana.
- The study findings demonstrate that healthcare professionals in this part of Ghana use mHealth applications to treat HIV, TB, hypertension, diabetes, and malaria conditions.
- The study results show a low-level use of mHealth applications for disease diagnosis and treatment support by healthcare professionals at the rural primary healthcare clinics.

*Recommendations for policy:*

- Our study encourages policymakers to deliberately implement mHealth technologies and applications at rural primary health clinics to support disease diagnosis and treatment procedures of patients’ conditions.
- Our study recommends that more primary studies be conducted focused on using mHealth interventions to treat and manage many diseases such as cancer, stroke, chronic respiratory conditions, asthma, and others in this region.
- The study encourages healthcare professionals to use mHealth applications to screen or diagnose several diseases such as neglected tropical diseases to enhance early detection.

## Introduction

Sub-Saharan African (SSA) counties, including Ghana, are confronted with a double burden of communicable and non-communicable diseases (1, 2). They also have weak healthcare systems, which has been exacerbated by the Severe acute respiratory syndrome coronavirus 2 (SARS-CoV-2) pandemic (1, 3-5). In addition, poor access to healthcare due to insufficient healthcare infrastructure, poor road networks, long-distance travel to health facilities, inadequate health education, lack of financial resources, insufficiently trained health professionals, and many others also further weakens the healthcare systems (6, 7). The government of Ghana (GoG) has committed to improving the digitization of healthcare systems, training and posting many skilled health professionals to rural communities, expanding mobile networks to rural Ghana (8).

Digitization of healthcare systems such as mobile health (mHealth) technologies and applications have been identified as promising strategies for improving access to healthcare delivery and patient outcomes (9, 10). Mobile health technology is defined as the use of portable devices for medical purposes (11). These applications have been shown to provide a cost-effective, convenient, and broadly accessible modality to implement population-level health interventions (12). In Ghana, mobile phones’ availability and utilization as of 2018 was reported to be about 52% and is expected to increase steadily (13). The high rate of mobile phone penetration and its innovativeness could become a promising tool to enhance healthcare provision and bridge the inequalities of healthcare accessibility (14-16). Mobile phone adoption and acceptability are disproportionately high in resource-limited settings (17). Thus, mHealth applications can address some healthcare disparities among hard-to-reach populations to help achieve universal health for all (18).

Studies in some low-and-middle-income countries (LMICs) have indicated that in this era of SARS-CoV-2, digital health technologies such as mHealth applications have been utilized for screening, diagnosis, risk assessment, tracking of real-time transmissions, and others in all settings (19-23). The use of mHealth applications has been shown to reduce the spread of SARS-CoV-2 and other infectious diseases in overcrowded emergency rooms and improved patient care (24-28). mHealth applications have also been deployed to support disease surveillance, medication, and treatment adherence, improve communication between clinical staff and their patients, appointment reminders, among others (29-33).

Despite these significant challenges and the limited resources in Ghana, mHealth interventions’ potential in playing a massive transformative role in healthcare provision has received considerable attention (11, 34). Considering the prospects of mHealth applications in resource-limited settings, we conducted a cross-sectional study to determine the availability and use of mHealth applications for disease diagnosis and treatment support by health workers in the Ashanti Region. Ghana. The research focuses on the availability of mHealth infrastructure, clinical staff competence, mHealth for diagnostics and treatment, usefulness, ease of use, user satisfaction, and behavioural intention to use mHealth applications. It is envisaged that the findings of this study will be beneficial to the GoG, donors, Non-Governmental Organizations in health, development partners, and others on improving quality healthcare provision by integrating mHealth applications into the normal clinical flow. It is also anticipated that our findings will assist the GoG and other similar settings to implement and sustain digital technologies such as mHealth to promote universal health coverage.

## Methods

### Study design and participants

A cross-sectional survey was conducted in primary healthcare facilities in the Ashanti Region of Ghana. The researchers conducted this survey to examine the availability and use of mHealth applications for disease diagnosis and treatment support by health professionals in the Ashanti Region of Ghana. In this survey, the participants are healthcare professionals who are highly trained clinical staff such as clinicians, nurses, laboratory scientists, pharmacists, physiotherapists, radiologists, and others who have been mandated to provide healthcare services to the public. Healthcare professionals across the one hundred health facilities gave written consent to take part in this survey. Few of the participants were assisted in answering the questionnaire, while the majority answered them independently. All the participants were working in healthcare facilities in the Ashanti Region of Ghana during our survey

### Study setting

The Ashanti Region is located in the middle part of Ghana (Figure 1). According to the 2010 population census, the Ashanti Region has over 4.70 million inhabitants with a growth rate of 2.7% and is described as Ghana’s business hub (35). It is projected to reach 9.5 million inhabitants in 2040, according to the Ghana Statistical Service 2012 report (36). This region is the most populated part of Ghana and has one of the enormous numbers of healthcare facilities in the entire country (37). This region is one of the areas with a high prevalence of several communicable and non-communicable diseases in Ghana. For instance, it has the second-highest prevalence rate of non-communicable diseases such as hypertension, stroke, diabetes, cancer, and others in Ghana (38-41). However, this region is the second most populated region in Ghana, yet it has one of the lowest tuberculosis prevalence rates (42). Ashanti Region is one of Ghana’s numerous areas with poor healthcare access, especially for people living in poor-resource settings. There are relatively moderate levels of accessibility to general primary healthcare; accessibility to healthcare services remains deficient in several rural districts in the Ashanti Region (43, 44). This is primarily due to the uneven distribution of healthcare facilities since most healthcare facilities are concentrated in the urban and semi-urban areas, with few rural communities (45).

**Figure 1:**
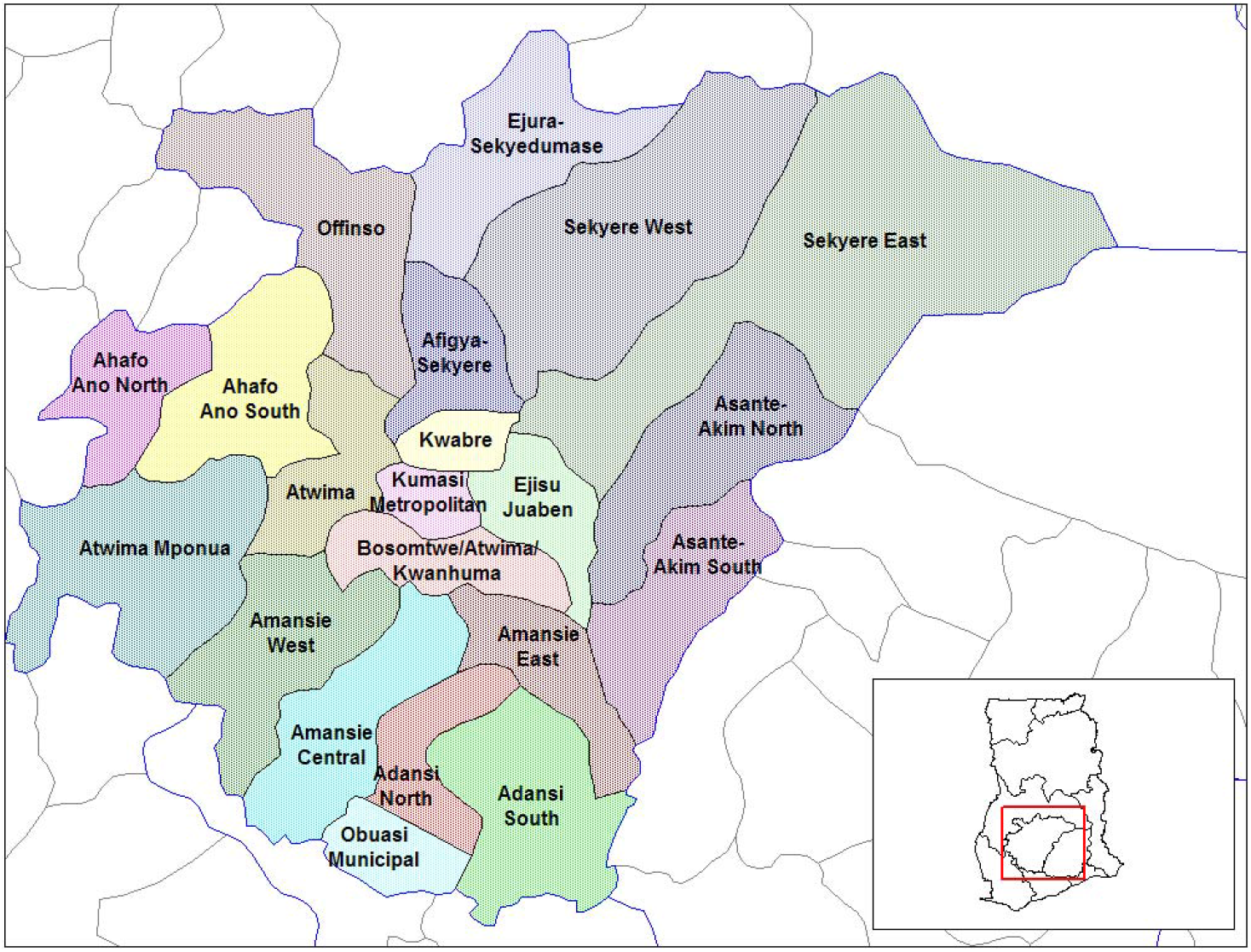
Map of Ashanti Region of Ghana (46)

### Sampling method

We obtained a list of 530 primary healthcare facilities from the Ashanti Regional Health Directorate (RHD) of the Ghana Health Service (GHS) (37). The researchers randomly selected 100 primary healthcare facilities from all the 43 districts in the Ashanti Region. Due to many healthcare facilities across the entire region, 100 healthcare facilities were chosen to ensure comprehensive study coverage. To guarantee the uniformity of sampled primary healthcare facilities in all the 43 districts, the following approach was employed: The primary healthcare facilities were first stratified into 43 strata, and each stratum denoting a district in the region. The 530 primary healthcare facilities were then grouped into four categories: 167 health centers, 154 clinics, 180 sub-district hospitals, and 29 district hospitals. Probability proportionate to size (PPS) was then used to determine the proportion of healthcare facilities from each stratum and category with the formula: nh = (Nh **/** N) x n where: nh represents the sample size for each stratum h; Nh represents population size for each stratum h; N representing the total population; and n denotes the total sample size. A purposive sampling technique was used to select all the district hospitals. Based on this, 29 hospitals were selected from category one, 30 clinics from category two, 28 clinics sampled from category three, and 13 chosen clinics from category four. We also used a proportionate stratification to get the total number of primary healthcare facilities selected from all the four groups in each of the 43 strata. After that, a simple random sampling technique was finally employed to select all the 100 healthcare facilities for this study (Additional file 1).

### Data collection and instruments

The researchers adopted the survey tool from studies conducted by Bauer et al. 2014, Bauer et al. 2017 and Abu-Dalbouch 2013 to match our study population, settings, and study aim (47-49). The cross-sectional survey tool (Additional file 2) was piloted in eight health centers and clinics in the Ashanti Region and modified to suit the local context based on the respondents’ feedback. This pilot study was conducted to ensure the validity, reliability, precision of data and remove all forms of ambiguity from the survey tool. We collected data on the category of health professionals, type of facility, number of healthcare professionals, number of patients seen per week, available healthcare infrastructure, healthcare workforce competence, ownership of mobile wireless devices, the usefulness of mHealth, ease of use of mHealth applications, user satisfaction, and behavioural intention to use mHealth applications. Data were surveyed and collected by the researcher and three trained research assistants.

### Ethics

This study was given full ethical clearance from the Biomedical Research Ethics Committee from the University of KwaZulu-Natal (approval reference no. BREC/00000202/2019), Ghana Health Service Ethics Review Committee (approval reference no. GHS-ERC006/11/19), regional clearance and recruitment site clearance of participants were obtained before the data collection commenced. All the study participants were given written consent forms explaining the study’s objective, and each signed the informed consent forms prior to their participation.

### Outcome measures

The study focused on examining the availability of mHealth technologies for disease diagnosis and treatment support by health professionals in the Ashanti Region of Ghana. The analysis of this study examined two outcome measures.

The first outcome was the availability of mobile health applications for disease diagnosis and treatment support which stemmed from the question: “Are there mHealth interventions available in this facility to support healthcare delivery?” A binary response (yes/no) was captured.

The second outcome was the use of mHealth applications for disease diagnosis and treatment support, which stemmed from the question: “What do you use mHealth interventions for” Responses were captured on four options: find health or medical information, disease diagnosis, treat and manage disease conditions, and treat and monitor patients’ health conditions.

### Explanatory variables

- Demographics assessed whether age, sex, categories of health professionals, type of health facility, the total number of healthcare professionals, and the number of patients who visit the facility per week influenced the availability and use of mHealth applications.
- Availability of health infrastructure assessed whether health infrastructure availability had any influence or facilitated the availability of mobile health applications for diagnostics and treatment support.
- Healthcare workforce competency assessed whether their level of knowledge influenced or facilitated the availability and use of mHealth applications for disease diagnosis and treatment support.
- Owning a mobile phone or having a mobile phone assessed whether mobile phone ownership had any influence or facilitated the use of mHealth applications for diagnostics and treatment support.
- The usefulness of mHealth applications assessed whether the benefits of mHealth applications had any influence or facilitated mHealth for diagnostics and treatment support.
- Ease of Use of mHealth applications assessed whether the easiness of using mHealth applications had any influence or facilitated mHealth for diagnostics and treatment support.
- User satisfaction of mHealth applications assessed whether the user satisfaction had any influence or facilitated mHealth for diagnostics and treatment support.
- Behavioural intention to use mHealth applications assessed whether behavioural intention to use mHealth had any influence or facilitated mHealth for diagnostics and treatment support.

### Data Management and analysis

The completed questionnaires were screened and reviewed by the principal investigator to complete and correct all discrepancies. Data were then captured into a passworded excel spreadsheet. After data cleaning and verification, the data set were then exported into STATA version 15. Descriptive statistics such as frequencies, percentages, means, and standard deviations characterize health workers’ demographics and clinical features. They were then presented in graphs, tables, and others. Multivariate logistic regression was employed to explore the influence of the demographic factors on the availability of mHealth for disease diagnosis and treatment support by healthcare workers. Again, this multivariate logistic regression was also used to explore the influence of the demographic factors on the use of mHealth for disease diagnosis and treatment support by health workers. In the multivariate logistic regression model, a p-value of 0.05 was set as the statistical significance. Furthermore, the associations were examined using the odds ratio and 95% CI estimates. A Chi-square test at a significance level of a p-value of 0.05 was used to assess the relationship between the availability and the use of mHealth for disease diagnosis and treatment support.

## Results

### Characteristics of the study participants

This study received a 100% response rate from the healthcare professionals in the selected healthcare facilities in the region. Completed responses were from 285 participants, with 146 males (51.23%) and 139 females (48.77%). The results reveal the highest participants were from those between the ages of 31-40 years, with 48.07%, followed by those in the category of 20-30 years, with 42.11% of the responses. However, the lowest participants were from 41-50 years and 51-60 years, with 9.47% and 0.35%. A majority (28.7%) of the respondents in this survey were general nurses, while only 2.46% were midwives. The average total number of health professionals in each healthcare facility was estimated at 57.8 (95% CI: 20-98). The average number of patients per week seen by these healthcare professionals was 175.4 (95% CI: 74-372) (Table S1 supplementary material file).

### Availability of mobile health for diagnostics and treatment support in the Ashanti region

Results from the frequency table (Table S 2 supplementary file) show that mobile wireless devices are available primarily to healthcare professionals with a frequency of 276 (96.84%). Mobile health applications are available with an estimated frequency of 179 (62.81%) and a non-availability frequency of 106 (37.19%). It is also clear that phone calls are the most predominant mHealth interventions utilized by healthcare professionals, with an estimated frequency of 183 (98.92%). Short message service (SMS) is another mHealth intervention used by healthcare professionals with a second higher frequency of 149 (80.54%). Figure 2 illustrates the availability of the various mHealth applications. Again, simple mobile phones are readily available to healthcare professionals with an estimated frequency of 185 (100%), followed by smartphones with 133 (71.89%) and tablets with 107 (57.84%). It is also observed that there is a higher continuous supply of electric power with an estimated frequency of 149 (80.54%) and relatively high available support systems of 106 (57.30%). Furthermore, most healthcare professionals have the requisite skills for diagnostics with a high frequency of 132 (71.36%) and the competence for treatment procedures with an estimated frequency of 164 (88.65%).

**Figure 2:**
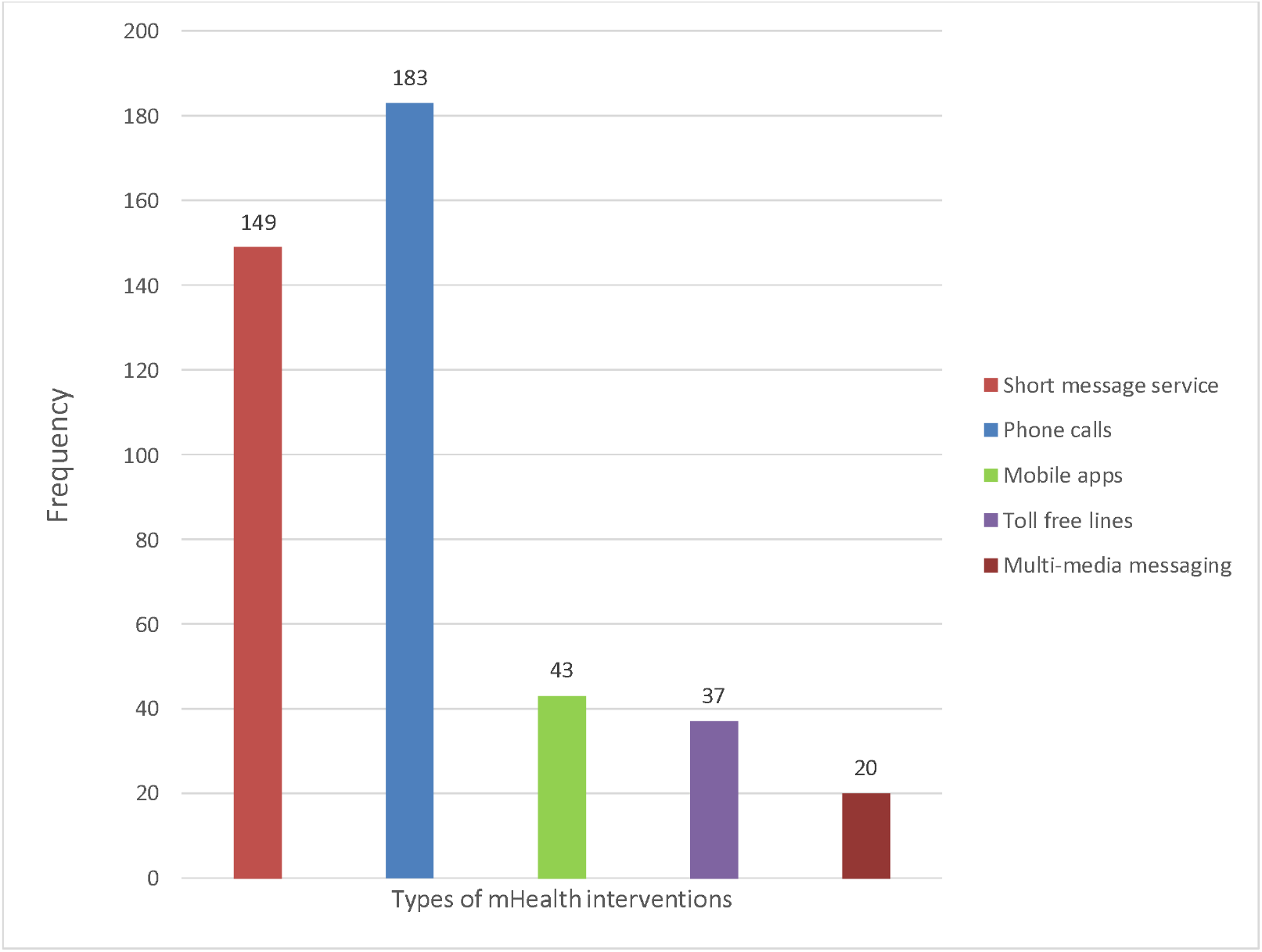
Availability of the various mHealth applications

### Use of mobile health for diagnostics and treatment support in the Ashanti region

The frequency table (Table S 3 supplementary material file) shows that the higher frequency rates of 182 (98.38%) indicate that many healthcare professionals are currently using mHealth applications to promote healthcare delivery. In this region, healthcare professionals use mHealth applications to support treatment procedures of diseases such as HIV (177), 95.86%, TB (171) 92.43%, hypertension 99 (53.51%), malaria 93 (50.54%), and diabetes 79 (42.70%). Figure 3 demonstrates various diseases that are being treated and managed with mHealth applications. However, only a few healthcare professionals use mHealth applications to support the treatment of other conditions such as diarrhoea 17 (9.19%), cancer 5 (2.70%), chronic respiratory disease 2 (1.08%), and stroke 0 (100%). Also, most healthcare professionals use mHealth applications to search for medical information 117 (63.24%), disease diagnosis 182 (98.38%), treat and manage disease conditions 162 (87.57%) and, treat and monitor patients’ health conditions 144 (77.84%).

**Figure 3:**
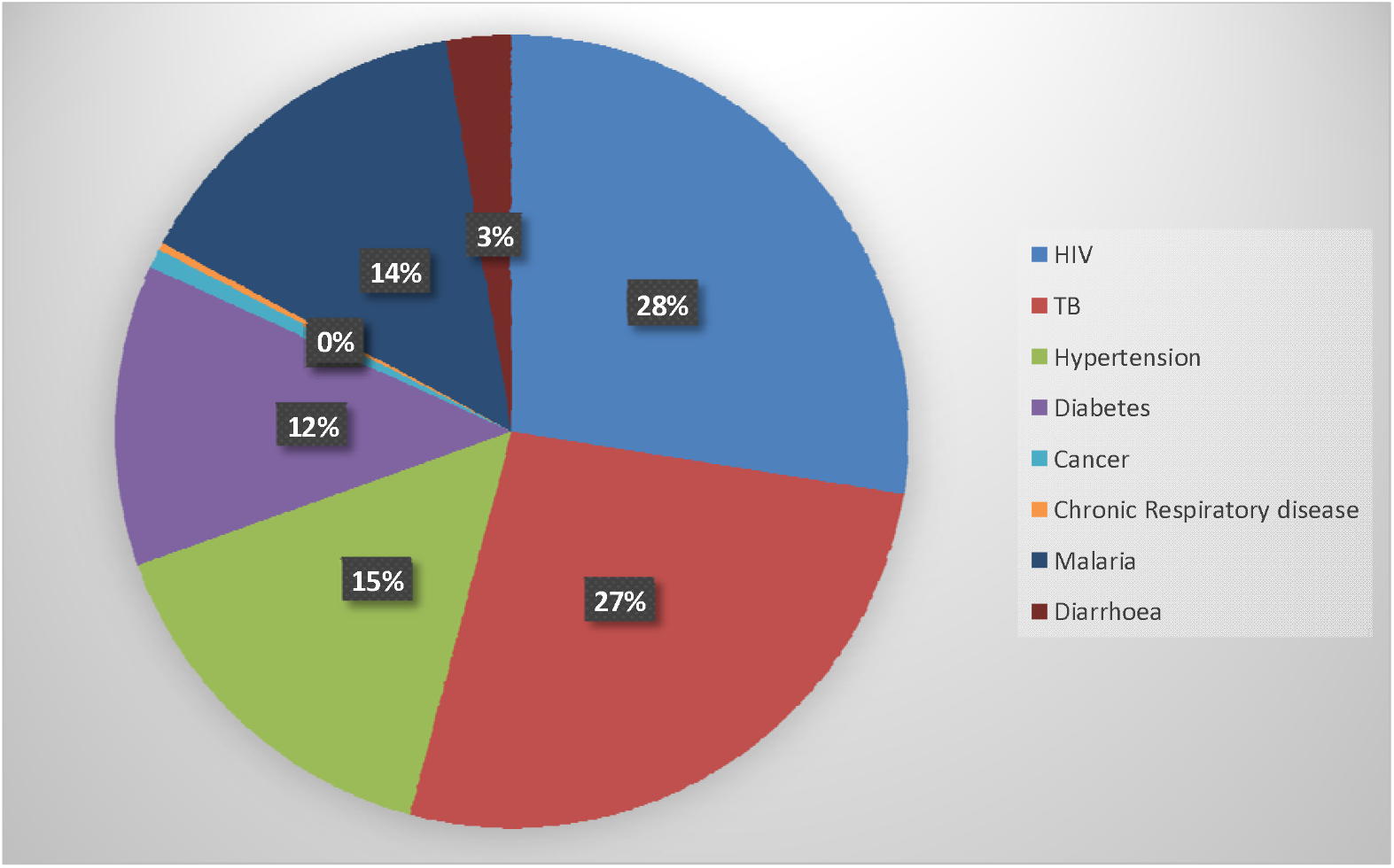
Types of diseases treated and managed with mHealth applications

Most healthcare professionals agreed that mHealth applications are easy to use when providing healthcare services to their clients. Majority of them confirmed that mHealth applications are easy to use to support disease diagnosis with an estimated frequency of 262 (87.37%). Some other healthcare professionals also indicated that it is flexible to interact with mHealth applications with an estimated frequency of 273 (95.79%). The survey reveals that healthcare professionals are comfortable using mHealth to support healthcare delivery with an estimated 266 (93.33%). Also, others are very confident in using mHealth applications with an estimated frequency of 254 (89.12%). Again, some healthcare professionals are delighted with the use of mHealth applications with an estimated frequency of 218 (76.49%). Moreover, most healthcare professionals would use mHealth to treat and manage patients’ disease conditions with a frequency of 254 (89.12%). Furthermore, others intend to use mHealth for disease diagnosis and treatment support with an estimated frequency of 279 (97.89%).

### Availability of health infrastructure and healthcare workforce competency

From the multivariate logistic regression model (Table 1), the results illustrate that healthcare workers within the age group 20-30 [OR = 17.8 (95% CI: 1.49-21.0)] and 31-40 [OR = 17.6 (95% CI: 1.45-21.1)] had increased odds for toll-free intervention availability when compared to healthcare workers above 40 years. Also, healthcare workers within the age group 20-30 and 31-40 had increased odds for mobile apps [OR = 1.46 (95% CI: 0.34-0.18)] and mHealth availability [OR = 2.93 (95% CI: 0.70-12.2)] compared to those above 40 years. Male healthcare workers had increased odds for mobile apps’ availability [OR = 1.27 (95% CI: 0.53-3.04)], mobile wireless devices [OR = 1.26 (95% CI: 0.11-5.16)] and toll-free intervention [OR = 1.02 (95% CI: 0.43-2.41)] compared to female healthcare workers (Figure 4).

**Table 1:** Multivariate analysis results for the availability of health infrastructure and healthcare workforce competency

| Variable |  | Mobile wireless devices |  |  | mHealth availability |  |  | SMS |  |  | Mobile apps |  |  | Toll-free |  |  |  |  |  |
| --- | --- | --- | --- | --- | --- | --- | --- | --- | --- | --- | --- | --- | --- | --- | --- | --- | --- | --- | --- |
|  |  | Odds ratio | 95% CI | p-value | Odds ratio | 95% CI | p-value | Odds ratio | 95% CI | p-value | Odds ratio | 95% CI | p-value | Odds ratio | 95% CI | p-value |  |  |  |
| Age | 20-30 years | 0.040 | 0.01-19.6 | 0.733 | 3.33 | 0.82-14.2 | 0.104 | 0.66 | 0.13-3.31 | 0.622 | 1.46** | 0.34-6.18 | 0.010 | 17.8** | 1.49-21.0 | 0.02 |  |  |  |
|  | 31-40 years | 0.020 | 0.03-78.8 | 0.648 | 2.93** | 0.70-12.2 | 0.010 | 1.49 | 0.29-7.68 | 0.632 | 2.21 | 0.51-9.64 | 0.287 | 17.6** | 1.45-21.1 | 0.02 |  |  |  |
|  | Above 40 years | 1 |  |  | 1 |  |  | 1 |  |  | 1 |  |  | 1 |  |  |  |  |  |
| Sex | Male | 1.260** | 0.11-5.16 | 0.02 | 0.40 | 0.18-0.85 | 0.018 | 0.81 | 0.34-1.96 | 0.653 | 1.27** | 0.53-3.04 | 0.010 | 1.02** | 0.43-2.41 | 0.002 |  |  |  |
|  | Female | 1 |  |  | 1 |  |  | 1 |  |  | 1 |  |  | 1 |  |  |  |  |  |
| Categories of health professionals | Doctors | - |  |  | 0.68 | 0.23-2.03 | 0.500 | 1.00 | 0.34-2.95 | 0.992 | 1.16 | 0.40-3.31 | 0.781 | 1.48 | 0.53-4.15 | 0.44 |  |  |  |
|  | Nurses | 6.980 | 0.66-73.5 | 0.106 | 0.67 | 0.23-1.91 | 0.458 | 1.52 | 0.48-4.85 | 0.471 | 0.84 | 0.27-2.55 | 0.764 | 0.96 | 0.33-2.83 | 0.95 |  |  |  |
|  | Pharmacists | 1.160 | 0.27-6.06 | 0.843 | 0.68 | 0.28-1.61 | 0.385 | 1.27 | 0.39-4.15 | 0.688 | 0.83 | 0.31-2.22 | 0.720 | 0.84 | 0.35-2.02 | 0.71 |  |  |  |
|  | Laboratory Scientists | 1 |  |  | 1 |  |  | 1 |  |  | 1 |  |  | 1 |  |  |  |  |  |
| Type of healthcare facility | District hospital | 0.010 | 0.45-1.28 | 0.993 | 0.04 | 0.01-2.26 | 0.119 | 2.16 | 0.04-1.00 | 0.693 | - | - | - | 23.8 | 0 | 0.98 |  |  |  |
|  | Health center | 0.087 | 0.32-2.01 | 0.996 | 0.53 | 0.17-1.68 | 0.287 | 0.59 | 0.08-4.13 | 0.600 | - | - | - | 84.7 | 0 | 0.98 |  |  |  |
|  | Clinic | 1 |  |  | 1 |  |  | 1 |  | 1 | 1 |  |  | 1 |  |  |  |  |  |
| Total number of healthcare professionals |  | 1.140 | 0.98-1.35 | 0.089 | 1.09** | 1.03-1.17 | 0.004 | 1.01 | 0.97-1.07 | 0.470 | 1.16*** | 1.07-1.25 | <0.001 | 1.01 | 0.95-1.07 | 0.60 |  |  |  |
| Total number of patients per week |  | 1.070 | 1.02-1.13 | 0.004 | 1.02*** | 1.01-1.04 | <0.001 | 0.99 | 0.99-1.00 | 0.885 | 1.00** | 1.00-1.01 | 0.020 | 0.97*** | 0.96-0.98 | <0.001 |  |  |  |
| Variable |  | Smartphones |  |  | Tablets |  |  | Supply of power |  |  | Support systems |  |  | Requisite skills |  |  | Competence to use mHealth |  |  |
|  |  | Odds ratio | 95 % | p-value | Odds | 95% CI | p-value | Odds ratio | 95% CI | p-value | Odds ratio | 95 % | p-value | Odds ratio | 95 % | p-value | Odds ratio | 95 % | p-value |

|  |  |  | CI | e | ratio |  |  |  |  | e |  | CI | e |  | CI | e |  | CI | e |
| --- | --- | --- | --- | --- | --- | --- | --- | --- | --- | --- | --- | --- | --- | --- | --- | --- | --- | --- | --- |
| Age | 20-30 years | 0.729 | 0.20-2.64 | 0.631 | 1.418 | 0.44-4.54 | 0.556 | 0.978 | 0.24-3.96 | 0.976 | 0.654 | 0.21 - 2.05 | 0.468 | 0.298 | 0.07-1.21 | 0.090 | 0.595 | 0.63-5.61 | 0.651 |
|  | 31-40 years | 0.823 | 0.22-3.05 | 0.771 | 1.769 | 0.54-5.83 | 0.348 | 1.491 | 0.35-6.26 | 0.585 | 0.871 | 0.27 - 2.78 | 0.817 | 0.604 | 0.14-2.47 | 0.485 | 1.805 | 0.16-20.1 | 0.631 |
|  | Above 40 years | 1 |  |  | 1 |  |  | 1 |  |  | 1 |  |  | 1 |  |  | 1 |  |  |
| Sex | Male | 0.917 | 0.44-1.89 | 0.814 | 0.985 | 0.49-1.97 | 0.968 | 1.075 | 0.45-2.55 | 0.870 | 1.209 | 0.63 - 2.31 | 0.565 | 1.208 | 0.59-2.46 | 0.601 | 0.671 | 0.20-2.24 | 0.516 |
|  | Female | 1 |  |  | 1 |  |  | 1 |  |  | 1 |  |  | 1 |  |  | 1 |  |  |
| Categories of health professionals | Doctors | 1.132 | 0.47-2.71 | 0.779 | 1.242 | 0.54-2.87 | 0.612 | 0.386 | 0.12-1.15 | 0.090 | 1.143 | 0.52 - 2.52 | 0.740 | 1.065* | 0.45-2.55 | 0.003 | 1.153** | 0.27-4.88 | 0.004 |
|  | Nurses | 1.236 | 0.49-3.08 | 0.649 | 1.690 | 0.45-2.56 | 0.881 | 0.345 | 0.11-1.06 | 0.065 | 0.787 | 0.35 - 1.77 | 0.563 | 0.723 | 0.30-1.73 | 0.467 | 0.345 | 0.08-1.39 | 0.136 |
|  | Pharmacists | 0.924 | 0.39-2.17 | 0.857 | 1.280 | 0.56-2.94 | 0.561 | 0.487 | 0.18-1.29 | 0.149 | 0.792 | 0.36 - 1.72 | 0.557 | 1.243* | 0.56-2.71 | 0.010 | 0.654 | 0.26-1.61 | 0.358 |
|  | Laboratory Scientists | 1 |  |  | 1 |  |  | 1 |  |  | 1 |  |  | 1 |  |  | 1 |  |  |
| Type of healthcare facility | District hospital | 0.872 | 0 | 0.984 | 0.119 | 0.008-2.94 | 0.193 | 59.87** | 70.06-5117 | <0.001 | 159.7* | 4.51 - 5660 | 0.005 | 21.66 | 0.73-639 | 0.075 | 0.623** | 0.11-0.35 | <0.001 |
|  | Health center | 0.1333 | 0 | 0.987 | 0.409 | 0.59-2.81 | 0.364 | 53.53** | 5.45-525 | 0.001 | 10.68* | 1.05-108 | 0.045 | 2.777 | 0.43-17.8 | 0.282 | 0.630 | 0.04-9.74 | 0.741 |
|  | Clinic | 1 |  |  | 1 |  |  | 1 |  |  | 1 |  |  | 1 |  |  | 1 |  |  |
| Total number of healthcare professionals |  | 1.073* | 1.02-1.12 | 0.002 | 1.057 | 1.02-1.10 | 0.007 | 0.907** | 0.85-0.96 | 0.001 | 0.948* | 0.81 - 0.89 | 0.015 | 0.969 | 0.93-1.01 | 0.160 | 1.196** | 1.09-1.31 | <0.001 |
| Total number of patients per week |  | 0.997 | 0.99-1.00 | 0.413 | 1.000 | 0.99-1.001 | 0.953 | 0.997 | 0.99-1.00 | 0.418 | 1.002 | 0.99 - 1.00 | 0.315 | 0.998 | 0.99-1.00 | 0.447 | 1.019** | 1.01-1.03 | 0.001 |

**Figure 4:**
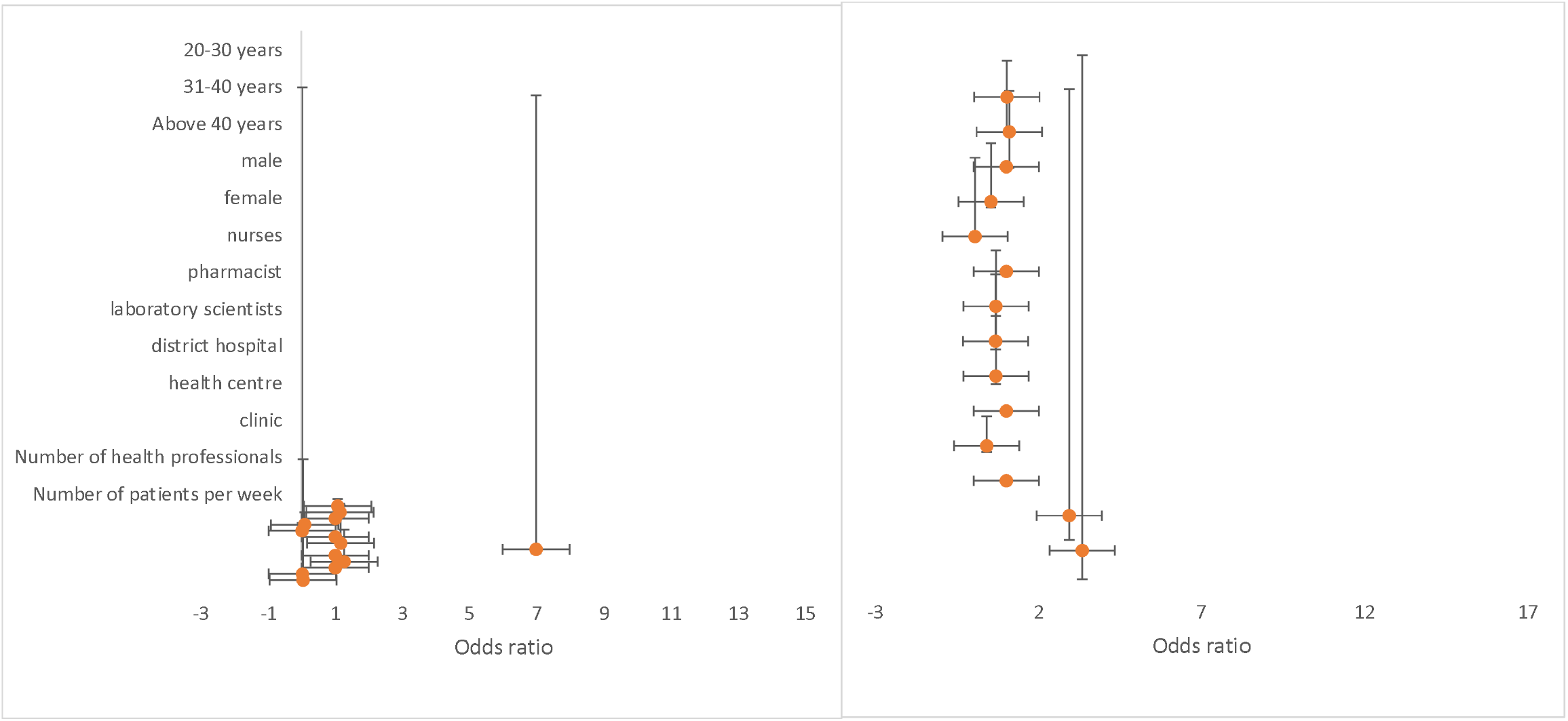
Odds ratio showing the association on the availability of mobile wireless devices and mHealth applications for disease diagnosis and treatment support by health workers in Ashanti region, Ghana.

Furthermore, the results show that health professionals such as doctors and pharmacists significantly influenced the requisite skills for diagnostics and competence to use mHealth for treatment support. Doctors had increased odds for the requisite skills for diagnostics and competence to use mHealth for treatment support [OR = 1.065 (95% CI: 0.45-2.55)] and [OR = 1.153 (95% CI: 0.27-4.88)] as compared to laboratory scientists. Pharmacists had increased odds for disease diagnosis requisite skills [OR = 1.243 (95% CI: 0.56-2.71)] compared to laboratory scientists. The results also illustrate those district hospitals and health centers significantly affect the supply of power and support systems.

Also, district hospitals increased the odds for the supply of power and support systems [OR = 59.87 (95% CI:70.06-5117)] and [OR =159.7 (95% CI: 4.51-5660)] compared to clinics. However, district hospitals had decreased odds [OR = 0.63 (95% CI: 0.11-0.35)] for the competence to use mHealth for treatment support. Health centers had increased odds for the supply of power and support systems [OR = 53.53 (95% CI: 5.45-525)] and [OR =10.68 (95% CI: 1.05-108)] compared to clinics. The total number of healthcare professionals with access to smartphones [OR = 1.073 (95% CI: 1.02-1.12)] and competence to use mHealth for treatment support [OR = 1.196 (95% CI: 1.09-1.31)] had increased odds. However, the total number of healthcare workers with access to supply of power [OR = 0.907 (95% CI: 0.85-0.96)] and support systems [OR = 0.948 (95% CI: 0.91-0.89)] had decreased in odds. Again, an increase in the number of patients per week increased odds for healthcare workers’ competence to use mHealth for treatment support [OR = 1.019 (95% CI: 1.01-1.03)] (Supplementary figure 1).

### Use of mHealth for diagnostics and treatment support

Results from the multivariate model (Table 2), healthcare workers within the age group 20-30 had increased odds for using mHealth to support the treatment of hypertension [OR = 2.28 (95% CI: 0.74-7.05)], diabetes [OR = 3.75 (95% CI: 0.96-14.6)], cancer [OR = 6.05 (95% CI: 0.01-5.85)], and malaria [OR = 1.04 (95% CI: 0.36-3.05)] compared to healthcare workers above 40 years. Also, healthcare workers within the age group 31-40 had increased odds for using mHealth for managing hypertension [OR = 2.12 (95% CI: 0.67-6.68)], diabetes [OR = 5.75 (95% CI: 1.43-23.1)], cancer [OR = 11.1 (95% CI: 0.01-12.0)] and malaria [OR = 1.24 (95% CI: 0.42-3.67)] as compared to healthcare workers above 40 years (Figure 5). Being a male healthcare professional raised the odds for the use mHealth to manage HIV [OR = 2.47 (95% CI: 0.37-16.4)] and TB [OR = 1.94 (95% CI: 0.49-7.62)] compared to being a female healthcare professional. Both medical doctors and nurses had increased odds [OR = 1.66 (95% CI: 0.30-9.16)] and [OR = 1.28 (95% CI: 0.28-5.83)] for the use of mHealth to manage TB when compared to laboratory scientists (Figure 5).

**Table 2:**
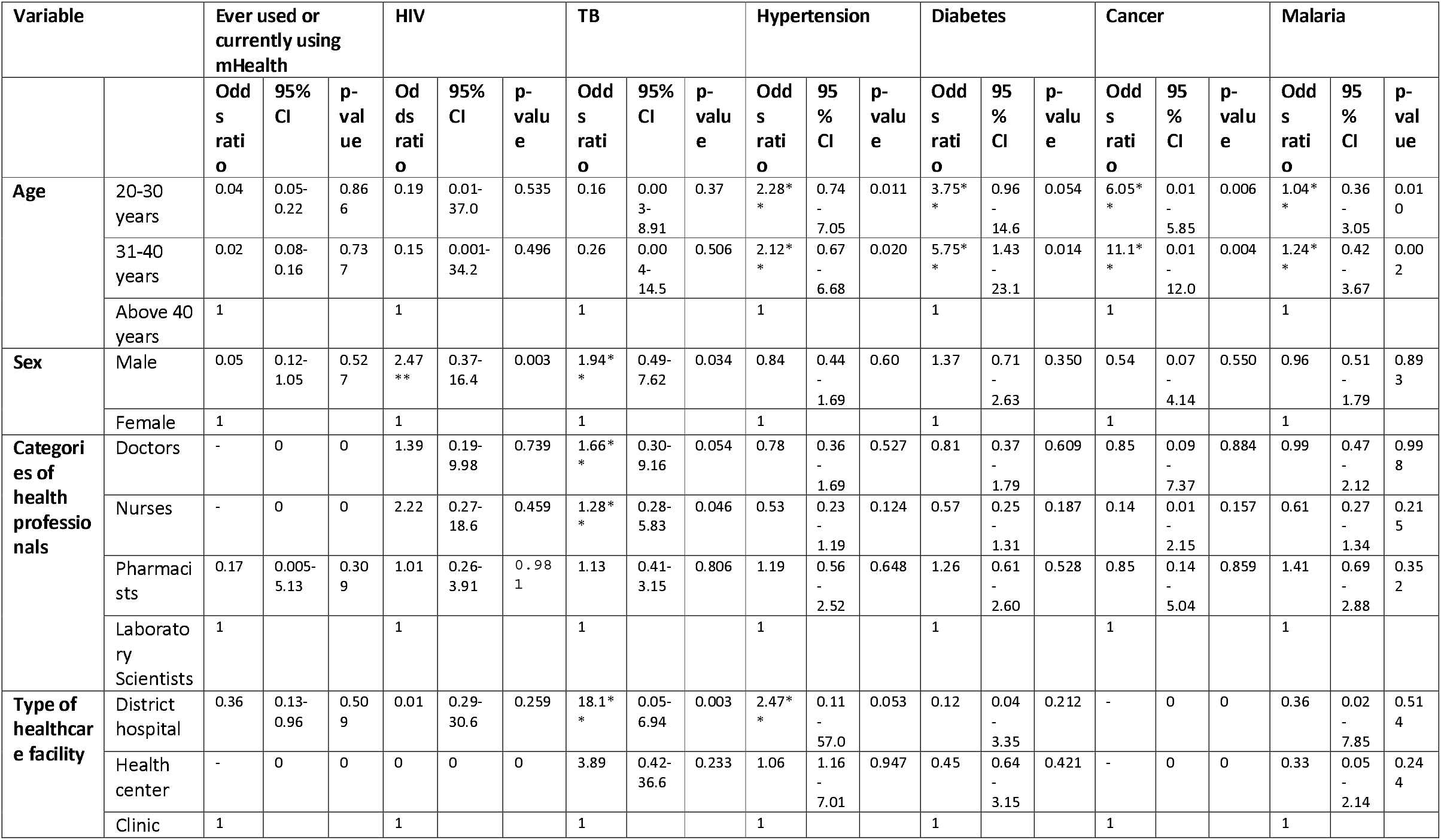
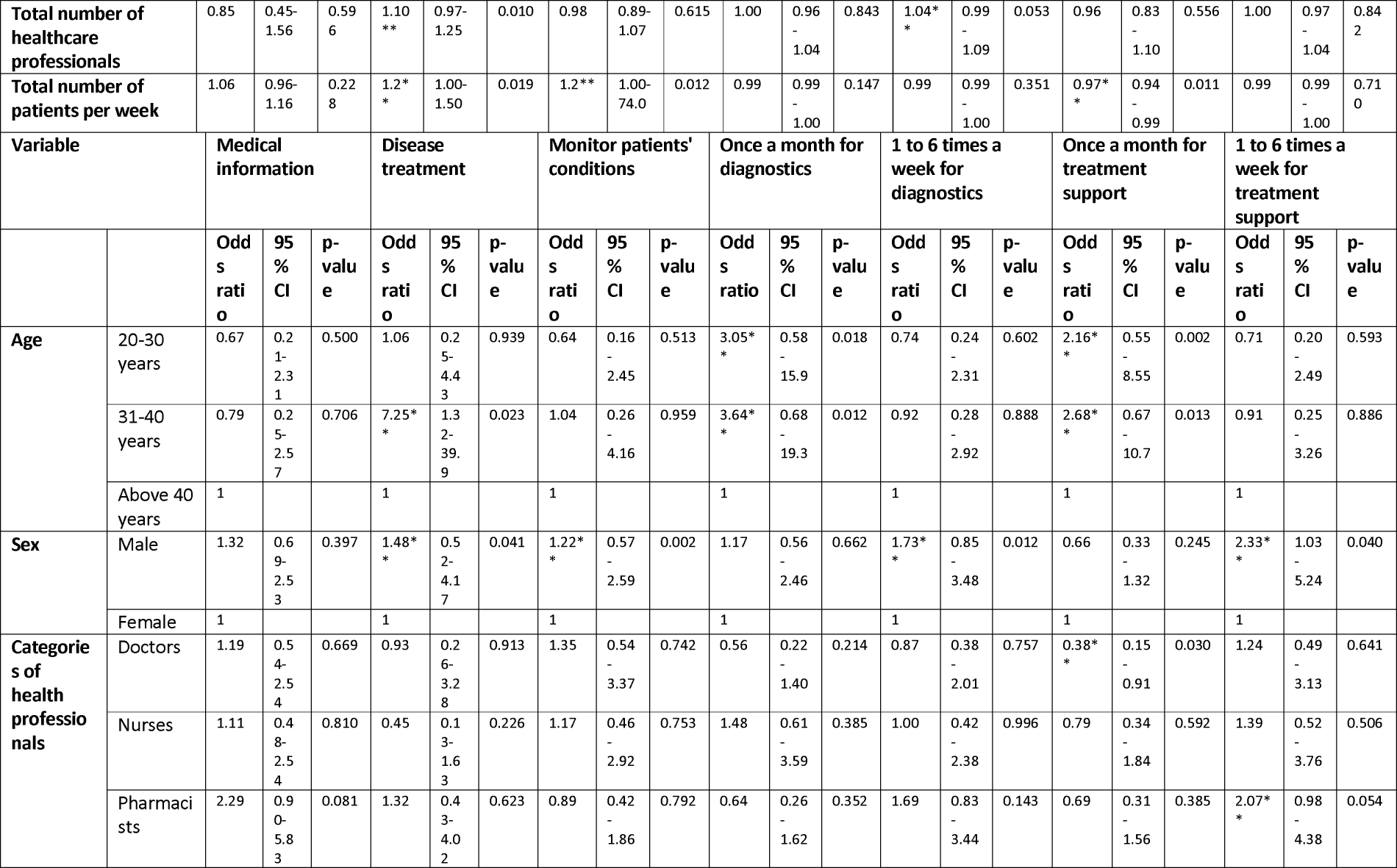
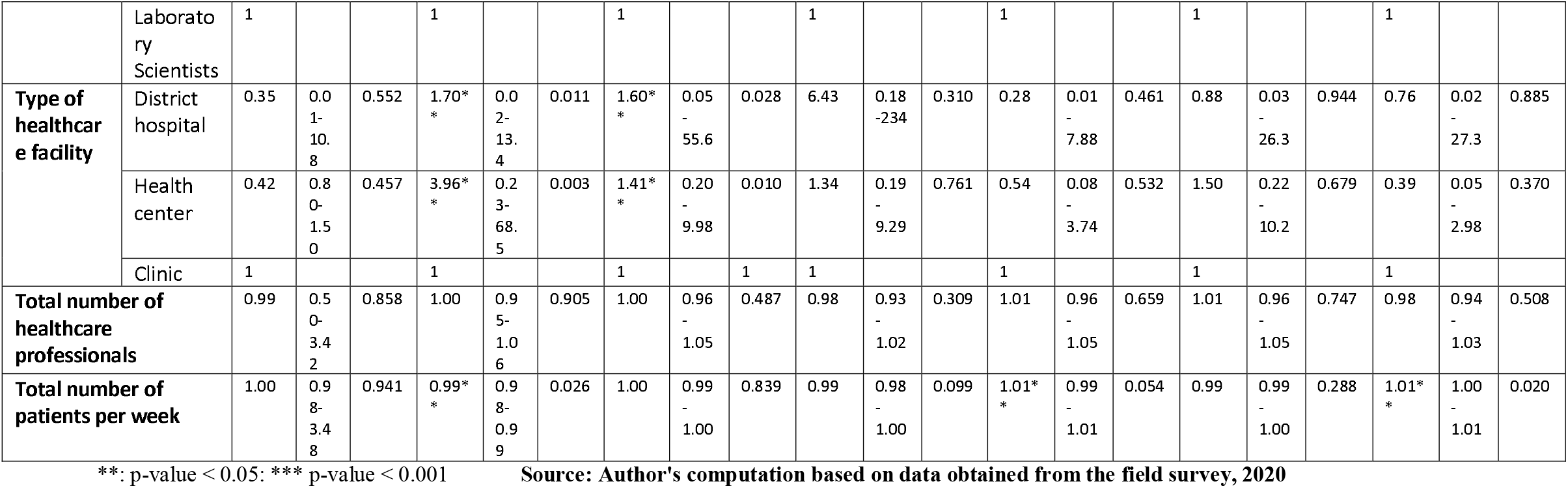
Multivariate analysis results for the use of mHealth for diagnostics and treatment support

**Figure 5:**
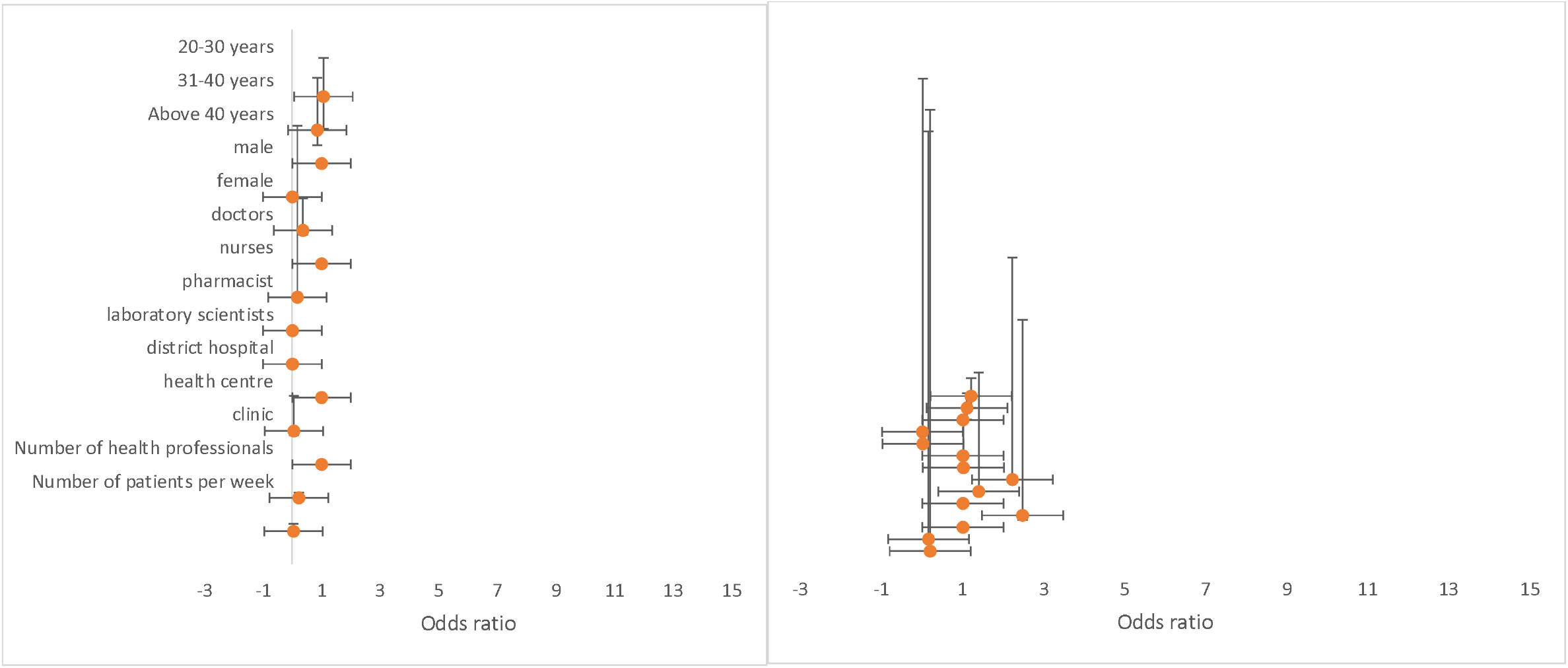
Odds ratio showing the association on the use of mHealth applications for the management and treatment of HIV and TB conditions by health workers in Ashanti region, Ghana

The results further show that healthcare workers within the age group 20-30 had increased odds for the use of mHealth for disease treatment [OR = 3.05 (95% CI: 0.58-15.9)], using mHealth once a month for diagnostics [OR = 2.16 (95% CI: 0.55-8.55)] and treatment support [OR = 1.06 (95% CI: 0.25-4.43)] compared to those above 40 years. Also, healthcare professionals within the age group 31-40 had a rise in odds for the use of mHealth for disease treatment [OR = 7.25 (95% CI: 1.32-39.9)], using mHealth once a month for diagnostics [OR = 3.64 (95% CI: 0.68-19.3)] and treatment support [OR = 2.68 (95% CI: 0.67-10.7)]. Being a male healthcare worker increased the odds for using mHealth to treat diseases [OR = 1.48 (95% CI: 0.52-4.17)], monitor patients’ conditions [OR = 1.22 (95% CI: 0.57-2.59)], using mHealth one to six times a week for diagnostics [OR = 1.73 (95% CI: 0.85-3.48)] and treatment support [OR = 2.33 (95% CI: 1.03-5.24)] compared to being a female healthcare worker.

Medical doctors had a decreased odds of using mHealth once a month for treatment support compared to laboratory scientists [OR = 0.38 (95% CI: 0.15-0.19)]. Again, pharmacists had increased odds for using mHealth application one to six times a week to support treatment [OR = 2.07 (95% CI: 0.98-4.35)] compared to laboratory scientists. District hospital increased the odds for the use of mHealth for disease treatment [OR = 1.70 (95% CI: 0.02-13.4)] and monitor patients’ conditions [OR = 1.60 (95% CI: 0.05-55.6)] compared to clinics. Also, health center had increased the odds for the use of mHealth for disease treatment [OR = 3.96 (95% CI: 0.23-68.5)] and monitor patients’ conditions [OR = 1.41 (95% CI: 0.20-9.98)] when to compared to clinics. As expected, a rise in the number of patients per week increased odds for using mHealth one to six times for diagnostics [OR = 1.01 (95% CI: 0.99-1.01)] and treatment support by healthcare workers [OR = 1.01 (95% CI: 1.00-1.01)]. However, an increase in the number of patients decreased the odds for using mHealth to treat diseases [OR = 0.99 (95% CI: 0.98-0.99)] (Supplementary file 2).

### Usefulness of mHealth interventions

The results from the multivariate model (Table 3) suggest that healthcare professionals within the age group 20-30 had reduced odds for the use of mHealth to monitor patients’ disease conditions [OR = 0.15 (95% CI: 0.02-1.07)], manage communicable diseases [OR = 0.15 (95% CI: 0.02-1.07)] and reminders for medication adherence [OR = 0.32 (95% CI: 0.08-1.24)] compared to those above 40 years. Also, healthcare workers within the age group 31-40 had increased odds for the use of mHealth to manage non-communicable diseases [OR = 1.23, (95% CI: 0.54-2.81)] and communicable diseases [OR = 1.41(95% CI: 0.54-3.82)] as compared to healthcare professionals above 40 years. However, healthcare professionals within the age group 31-40 had reduced odds for the use of mHealth as reminders for treatment adherence procedures [OR = 0.41(95% CI: 0.17-0.99)] when compared to those above 40 years. Male healthcare professionals who use mHealth to monitor patients’ disease conditions [OR = 1.76 (95% CI: 0.80-3.85)], manage communicable diseases [OR = 1.19 (95% CI: 0.72-2.00)], manage non-communicable diseases [OR = 1.19 (95% CI: 0.72-2.10)] and as reminders for medication adherence [OR = 1.31 (95% CI: 0.67-2.54)] had increased odds when compared to female healthcare professionals.

**Table 3:**
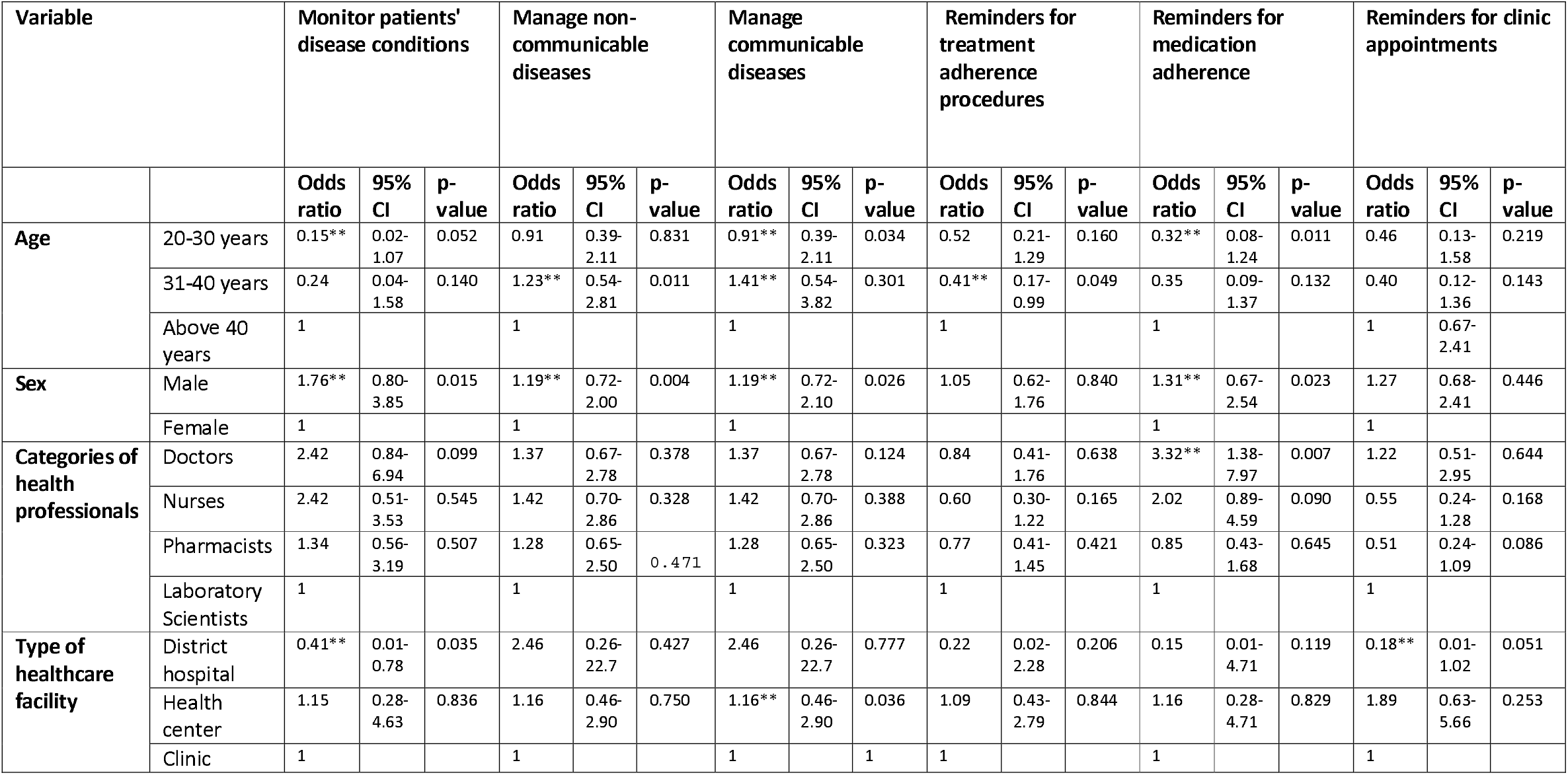
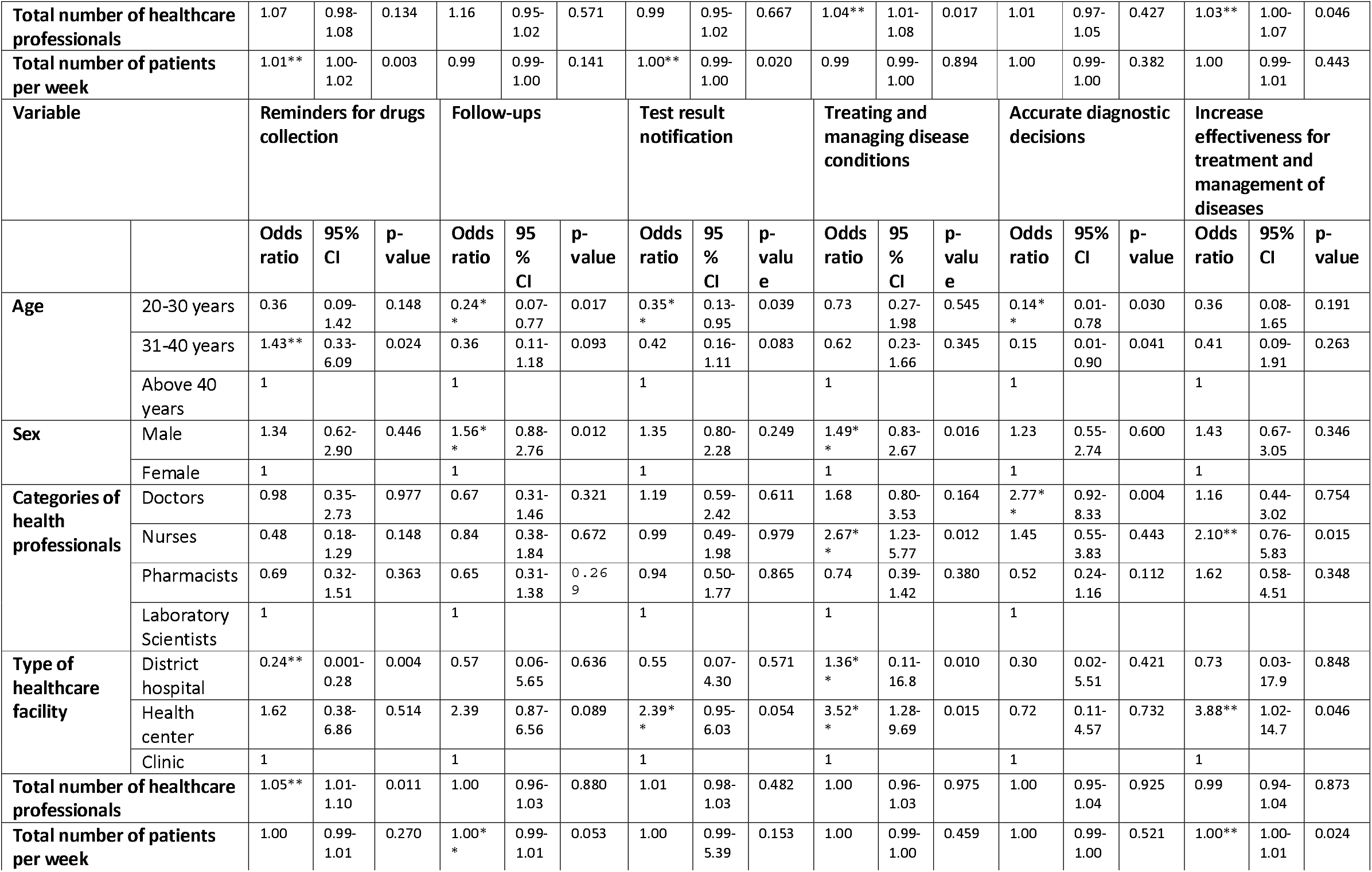
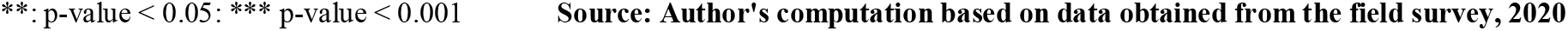
Multivariate analysis results for the usefulness of mHealth interventions

| Variable |  | Monitor patients' disease conditions |  |  | Manage non-communicable diseases |  |  | Manage communicable diseases |  |  | Reminders for treatment adherence procedures |  |  | Reminders for medication adherence |  |  | Reminders for clinic appointments |  |  |
| --- | --- | --- | --- | --- | --- | --- | --- | --- | --- | --- | --- | --- | --- | --- | --- | --- | --- | --- | --- |
|  |  | Odds ratio | 95% CI | p-value | Odds ratio | 95% CI | p-value | Odds ratio | 95% CI | p-value | Odds ratio | 95% CI | p-value | Odds ratio | 95% CI | p-value | Odds ratio | 95% CI | p-value |
| Age | 20-30 years | 0.15** | 0.02-1.07 | 0.052 | 0.91 | 0.39-2.11 | 0.831 | 0.91** | 0.39-2.11 | 0.034 | 0.52 | 0.21-1.29 | 0.160 | 0.32** | 0.08-1.24 | 0.011 | 0.46 | 0.13-1.58 | 0.219 |
|  | 31-40 years | 0.24 | 0.04-1.58 | 0.140 | 1.23** | 0.54-2.81 | 0.011 | 1.41** | 0.54-3.82 | 0.301 | 0.41** | 0.17-0.99 | 0.049 | 0.35 | 0.09-1.37 | 0.132 | 0.40 | 0.12-1.36 | 0.143 |
|  | Above 40 years | 1 |  |  | 1 |  |  | 1 |  |  | 1 |  |  | 1 |  |  | 1 | 0.67-2.41 |  |
| Sex | Male | 1.76** | 0.80-3.85 | 0.015 | 1.19** | 0.72-2.00 | 0.004 | 1.19** | 0.72-2.10 | 0.026 | 1.05 | 0.62-1.76 | 0.840 | 1.31** | 0.67-2.54 | 0.023 | 1.27 | 0.68-2.41 | 0.446 |
|  | Female | 1 |  |  | 1 |  |  | 1 |  |  |  |  |  | 1 |  |  | 1 |  |  |
| Categories of health professionals | Doctors | 2.42 | 0.84-6.94 | 0.099 | 1.37 | 0.67-2.78 | 0.378 | 1.37 | 0.67-2.78 | 0.124 | 0.84 | 0.41-1.76 | 0.638 | 3.32** | 1.38-7.97 | 0.007 | 1.22 | 0.51-2.95 | 0.644 |
|  | Nurses | 2.42 | 0.51-3.53 | 0.545 | 1.42 | 0.70-2.86 | 0.328 | 1.42 | 0.70-2.86 | 0.388 | 0.60 | 0.30-1.22 | 0.165 | 2.02 | 0.89-4.59 | 0.090 | 0.55 | 0.24-1.28 | 0.168 |
|  | Pharmacists | 1.34 | 0.56-3.19 | 0.507 | 1.28 | 0.65-2.50 | 0.471 | 1.28 | 0.65-2.50 | 0.323 | 0.77 | 0.41-1.45 | 0.421 | 0.85 | 0.43-1.68 | 0.645 | 0.51 | 0.24-1.09 | 0.086 |
|  | Laboratory Scientists | 1 |  |  | 1 |  |  | 1 |  |  | 1 |  |  | 1 |  |  | 1 |  |  |
| Type of healthcare facility | District hospital | 0.41** | 0.01-0.78 | 0.035 | 2.46 | 0.26-22.7 | 0.427 | 2.46 | 0.26-22.7 | 0.777 | 0.22 | 0.02-2.28 | 0.206 | 0.15 | 0.01-4.71 | 0.119 | 0.18** | 0.01-1.02 | 0.051 |
|  | Health center | 1.15 | 0.28-4.63 | 0.836 | 1.16 | 0.46-2.90 | 0.750 | 1.16** | 0.46-2.90 | 0.036 | 1.09 | 0.43-2.79 | 0.844 | 1.16 | 0.28-4.71 | 0.829 | 1.89 | 0.63-5.66 | 0.253 |
|  | Clinic | 1 |  |  | 1 |  |  | 1 |  | 1 | 1 |  |  | 1 |  |  | 1 |  |  |
| **Total number of healthcare professionals** |  | 1.07 | 0.98-1.08 | 0.134 | 1.16 | 0.95-1.02 | 0.571 | 0.99 | 0.95-1.02 | 0.667 | 1.04** | 1.01-1.08 | 0.017 | 1.01 | 0.97-1.05 | 0.427 | 1.03** | 1.00-1.07 | 0.046 |
| **Total number of patients per week** |  | 1.01** | 1.00-1.02 | 0.003 | 0.99 | 0.99-1.00 | 0.141 | 1.00** | 0.99-1.00 | 0.020 | 0.99 | 0.99-1.00 | 0.894 | 1.00 | 0.99-1.00 | 0.382 | 1.00 | 0.99-1.01 | 0.443 |
| **Variable** |  | **Reminders for drugs collection** |  |  | **Follow-ups** |  |  | **Test result notification** |  |  | **Treating and managing disease conditions** |  |  | **Accurate diagnostic decisions** |  |  | **Increase effectiveness for treatment and management of diseases** |  |  |
|  |  | **Odds ratio** | **95% CI** | **p-value** | **Odds ratio** | **95% CI** | **p-value** | **Odds ratio** | **95% CI** | **p-value** | **Odds ratio** | **95% CI** | **p-value** | **Odds ratio** | **95% CI** | **p-value** | **Odds ratio** | **95% CI** | **p-value** |
| **Age** | 20-30 years | 0.36 | 0.09-1.42 | 0.148 | 0.24* | 0.07-0.77 | 0.017 | 0.35* | 0.13-0.95 | 0.039 | 0.73 | 0.27-1.98 | 0.545 | 0.14* | 0.01-0.78 | 0.030 | 0.36 | 0.08-1.65 | 0.191 |
|  | 31-40 years | 1.43** | 0.33-6.09 | 0.024 | 0.36 | 0.11-1.18 | 0.093 | 0.42 | 0.16-1.11 | 0.083 | 0.62 | 0.23-1.66 | 0.345 | 0.15 | 0.01-0.90 | 0.041 | 0.41 | 0.09-1.91 | 0.263 |
|  | Above 40 years | 1 |  |  | 1 |  |  | 1 |  |  | 1 |  |  | 1 |  |  | 1 |  |  |
| **Sex** | Male | 1.34 | 0.62-2.90 | 0.446 | 1.56* | 0.88-2.76 | 0.012 | 1.35 | 0.80-2.28 | 0.249 | 1.49* | 0.83-2.67 | 0.016 | 1.23 | 0.55-2.74 | 0.600 | 1.43 | 0.67-3.05 | 0.346 |
|  | Female | 1 |  |  | 1 |  |  | 1 |  |  | 1 |  |  | 1 |  |  | 1 |  |  |
| **Categories of health professionals** | Doctors | 0.98 | 0.35-2.73 | 0.977 | 0.67 | 0.31-1.46 | 0.321 | 1.19 | 0.59-2.42 | 0.611 | 1.68 | 0.80-3.53 | 0.164 | 2.77* | 0.92-8.33 | 0.004 | 1.16 | 0.44-3.02 | 0.754 |
|  | Nurses | 0.48 | 0.18-1.29 | 0.148 | 0.84 | 0.38-1.84 | 0.672 | 0.99 | 0.49-1.98 | 0.979 | 2.67* | 1.23-5.77 | 0.012 | 1.45 | 0.55-3.83 | 0.443 | 2.10** | 0.76-5.83 | 0.015 |
|  | Pharmacists | 0.69 | 0.32-1.51 | 0.363 | 0.65 | 0.31-1.38 | 0.269 | 0.94 | 0.50-1.77 | 0.865 | 0.74 | 0.39-1.42 | 0.380 | 0.52 | 0.24-1.16 | 0.112 | 1.62 | 0.58-4.51 | 0.348 |
|  | Laboratory Scientists | 1 |  |  | 1 |  |  | 1 |  |  | 1 |  |  | 1 |  |  |  |  |  |
| **Type of healthcare facility** | District hospital | 0.24** | 0.001-0.28 | 0.004 | 0.57 | 0.06-5.65 | 0.636 | 0.55 | 0.07-4.30 | 0.571 | 1.36* | 0.11-16.8 | 0.010 | 0.30 | 0.02-5.51 | 0.421 | 0.73 | 0.03-17.9 | 0.848 |
|  | Health center | 1.62 | 0.38-6.86 | 0.514 | 2.39 | 0.87-6.56 | 0.089 | 2.39* | 0.95-6.03 | 0.054 | 3.52* | 1.28-9.69 | 0.015 | 0.72 | 0.11-4.57 | 0.732 | 3.88** | 1.02-14.7 | 0.046 |
|  | Clinic | 1 |  |  | 1 |  |  | 1 |  |  | 1 |  |  | 1 |  |  | 1 |  |  |
| **Total number of healthcare professionals** |  | 1.05** | 1.01-1.10 | 0.011 | 1.00 | 0.96-1.03 | 0.880 | 1.01 | 0.98-1.03 | 0.482 | 1.00 | 0.96-1.03 | 0.975 | 1.00 | 0.95-1.04 | 0.925 | 0.99 | 0.94-1.04 | 0.873 |
| **Total number of patients per week** |  | 1.00 | 0.99-1.01 | 0.270 | 1.00* | 0.99-1.01 | 0.053 | 1.00 | 0.99-5.39 | 0.153 | 1.00 | 0.99-1.00 | 0.459 | 1.00 | 0.99-1.00 | 0.521 | 1.00** | 1.00-1.01 | 0.024 |

\*\*: p-value < 0.05; \*\*\* p-value < 0.001
Source: Author's computation based on data obtained from the field survey, 2020

Medical doctors had 3-fold increased odds of using mHealth as reminders for medication adherence compared with laboratory scientists [OR = 3.32 (95% CI: 1.38-7.97)]. District hospital reduced the odds for the use of mHealth to monitor patients’ disease conditions [OR = 0.41 (95% CI: 0.01-0.78)] and as reminders for clinic appointments [OR = 0.18 (95% CI: 0.01-1.02)] when compared to clinics. Health center increased the odds for the use of mHealth to manage communicable diseases as compared to clinics [OR = 1.16 (95% CI: 0.46-2.90)]. The total number of healthcare professionals who use mHealth as reminders for treatment adherence procedures [OR = 1.04 (95% CI: 1.01-1.08)] and clinic appointments [OR = 1.03 (95% CI: 1.00-1.07)] had risen odds. A rise in number of patients per week increased odds for the use of mHealth to monitor patients’ disease conditions [OR = 1.01 (95% CI: 1.00-1.02)] and manage communicable diseases [OR = 1.00 (95% CI: 0.99-1.00)].

District hospitals increased the odds for mHealth to treat and manage disease conditions than clinics [OR = 1.36 (95% CI: 0.11-16.8)]. However, as a district hospital, the odds for the use of mHealth as reminders to collect drugs reduced [OR = 0.24 (95% CI: 0.001-0.28)]. Also, a health center increased the odds for the use of mHealth to notify patients of their test results [OR = 2.39 (95% CI: 0.95-6.03)], treat and manage disease conditions [OR = 3.52 (95% CI: 1.28-9.69)] and increase effectiveness for treatment and management of diseases [OR = 3.88 (95% CI: 1.02-14.7)] as compared to clinics. The total number of healthcare professionals who use mHealth as reminders for drug collection had increased odds [OR = 1.05 (95% CI: 1.01-1.10)]. An increase in the number of patients per week had increased the odds for the use of mHealth for follow-ups [OR = 1.00 (95% CI: 0.99-1.01)] and increase effectiveness to treat and manage diseases [OR = 1.00 (95% CI: 1.00-1.01)].

### Ease of Use of mHealth interventions

In the multivariate logistic regression model (Table 4), the results demonstrate that healthcare professionals within the age groups 20-30 and 31-40 had increased the odds for the flexibility to interact with mHealth devices [OR = 1.16 (95% CI: 0.11-11.8)] and easy to use mHealth for treatment support [OR = 1.33 (95% CI: 0.17-10.3)] compared to those above 40 years. Being a male healthcare worker increased the odds for easy-to-use mHealth for disease diagnosis [OR = 1.71 (95% CI: 0.67-4.29)] and the flexibility to interact with mHealth [OR = 4.00 (95% CI: 0.76-20.9)] compared to being female healthcare professional. Medical doctors had 9-fold increased odds of becoming skillful in using mHealth for disease diagnosis and treatment support [OR = 9.56 (95% CI: 1.78-51.1)] compared to laboratory scientists. Again, nurses had 2-fold increased odds for easy-to-use mHealth for disease diagnosis [OR = 2.66 (95% CI: 0.82-8.62)] when compared to laboratory scientists.

**Table 4:**
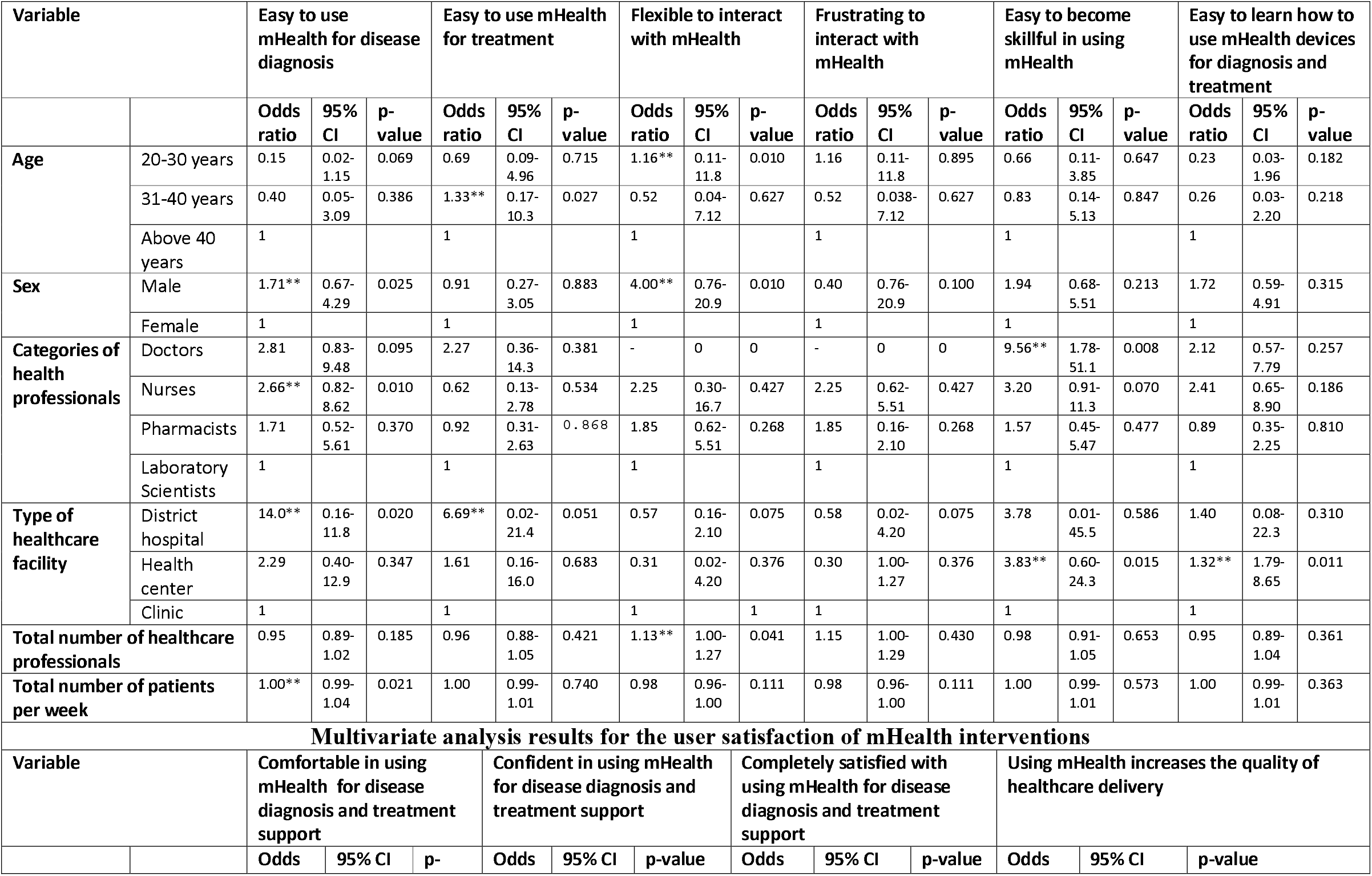
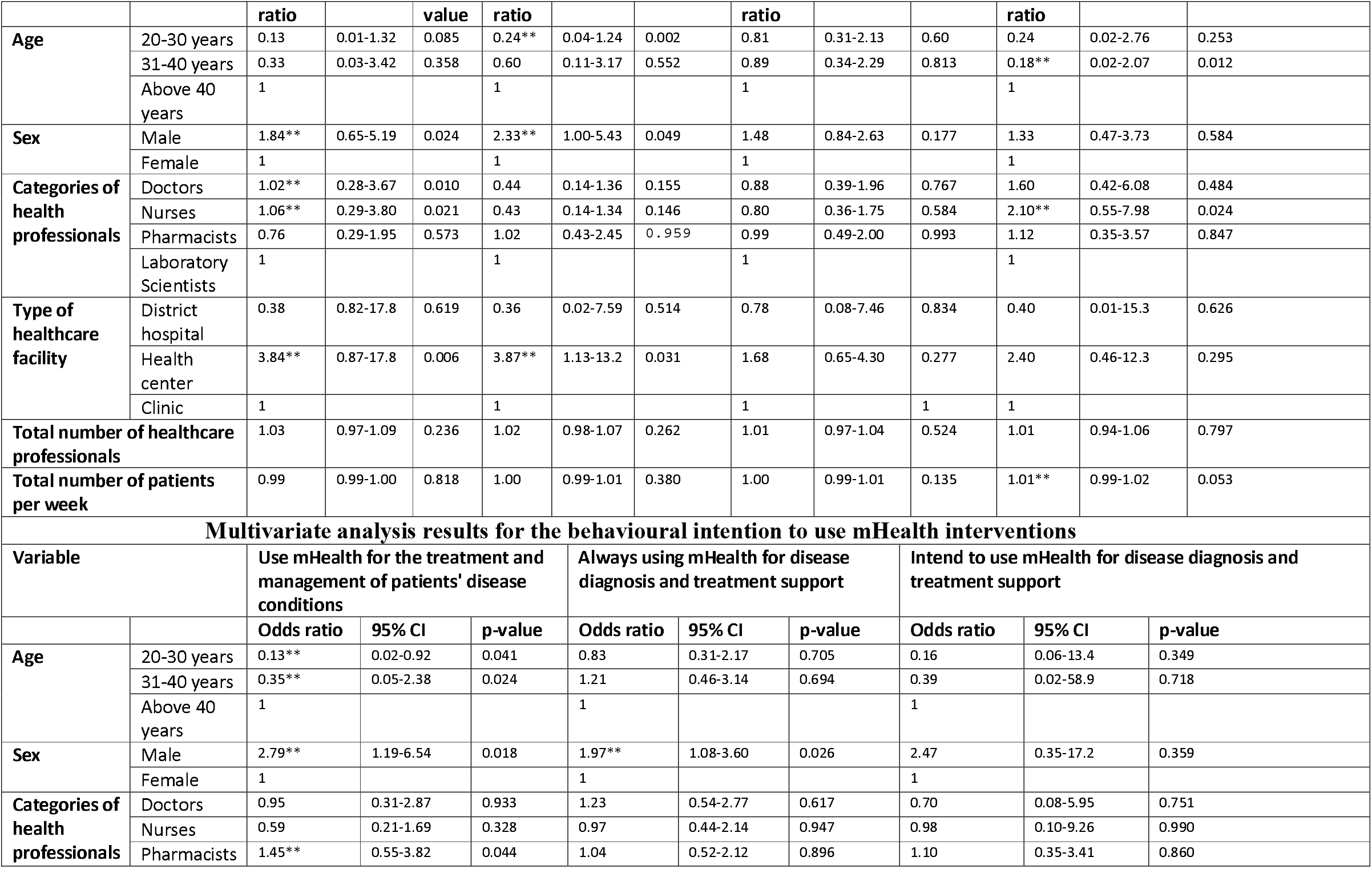
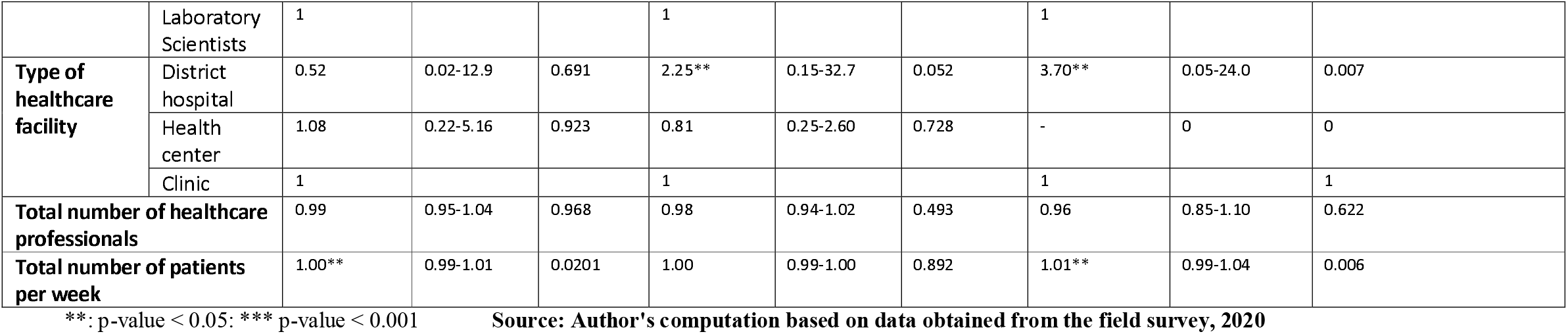
Multivariate analysis results for the ease of use of mHealth, user satisfaction of mHealth and behavioural intention to use mHealth interventions

Also, district hospital had increased the odds for easy-to-use mHealth for disease diagnosis [OR = 14.0 (95% CI: 0.16-11.8)] and treatment support [OR = 6.69 (95% CI: 0.02-21.4)] compared to clinics. A health center had increased odds for easy learning of how to use mHealth devices [OR = 1.32 (95% CI: 1.79-8.65)] and becoming skillful in using such applications for disease diagnosis and treatment support [OR = 1.32 (95% CI: 0.60-24.3)]. The total number of healthcare professionals had increased odds for the flexibility of interacting with mHealth devices for disease diagnosis and treatment support [OR = 1.13 (95% CI: 1.00-1.27)]. A rise in the number of patients per week had increased odds for using mHealth for disease diagnosis easily [OR = 1.00 (95% CI: 0.99-1.04)].

### User satisfaction of mHealth interventions

The results from the multivariate model (Table 4) show that healthcare workers within the age groups 20-30 and 31-40 had reduced odds for healthcare workers’ confidence in using mHealth for disease diagnosis and treatment support [OR = 0.24 (95% CI: 0.04-1.24)] and mHealth increasing the quality of healthcare delivery [OR = 0.18 (95% CI: 0.02-2.07)] compared to those above 40 years. Being a male healthcare professional increased the odds of healthcare workers’ comfortability [OR = 1.84 (95% CI: 0.65-5.19)] and confidence [OR = 2.33 (95% CI: 1.00-5.43)] in using mHealth for disease diagnosis and treatment support compared to being female healthcare professional.

Again, medical doctors had increased the odds of becoming comfortable using mHealth applications for disease diagnosis and treatment support [OR = 1.02 (95% CI: 0.28-3.80)] compared to being a laboratory scientist. Again, nurses had increased the odds of feeling comfortable with mHealth applications [OR = 1.06 (95% CI: 0.29-3.80)] and improving the quality of healthcare delivery with mHealth applications [OR = 2.10 (95% CI: 0.55-7.98)] when compared to being laboratory scientists. Health center had increased odds of healthcare workers’ comfortability [OR = 3.84 (95% CI: 0.87-17.8)] and confidence [OR = 3.87 (95% CI: 1.13-13.2)] with the use of mHealth for disease diagnosis and treatment support compared to clinics. An increase in the number of patients per week had increased the odds of using mHealth to improve healthcare delivery quality [OR = 1.01 (95% CI: 0.99-1.02)].

### Behavioural intention to use mHealth interventions

Results from the multivariate model (Table 4) reveal that healthcare professionals within the age groups 20-30 and 31-40 had increased odds for healthcare professionals intending to use mHealth for the treatment [OR = 0.13 (95% CI: 0.02-0.92)] and management of patients’ disease conditions [OR = 0.35 (95% CI: 0.05-2.38)] compared to those above 40 years. Being a male healthcare professional increased the odds of healthcare workers’ intention to use mHealth for treating and managing patients’ disease conditions [OR = 2.79 (95% CI: 1.19-6.54)] and disease diagnosis and treatment support [OR = 1.97 (95% CI: 1.08-3.60)] compared to being female healthcare professional.

Also, pharmacists had increased the odds of healthcare workers’ intention to use mHealth to treat and manage patients’ disease conditions compared to laboratory scientists [OR = 1.45 (95% CI: 0.55-3.82)]. The odds increased for a district hospital where healthcare workers intend to use mHealth [OR = 2.25 (95% CI: 0.15-32.7)] and will always use mHealth for disease diagnosis and treatment support [OR = 3.20 (95% CI: 0.05-24.0)] compared to clinics. A rise in the number of patients per week had increased the odds for healthcare workers using mHealth to treat and manage patients’ disease conditions [OR = 1.00 (95% CI: 0.99-1.01)] and their intention to use mHealth for disease diagnosis and treatment support [OR = 1.01 (95% CI: 0.99-1.04)].

### Association between health infrastructure availability or healthcare workforce competency and ownership of mobile wireless devices

A cross-sectional tabulation was done between healthcare infrastructure’s availability or healthcare workforce competency and ownership of mobile wireless devices using a chi-square test (Table S 4 supplementary material file). The chi-square test results illustrate a significant relationship between mobile wireless devices’ availability and currently using mHealth interventions to support healthcare provision (p< 0.05). Healthcare workers with mobile wireless devices were likely to use mHealth interventions to help healthcare delivery than those without mobile wireless devices. In addition to that, there is an association between mobile wireless devices’ availability and their use to assist malaria conditions’ treatment is statistically significant (p< 0.05). Healthcare workers with mobile wireless devices were more likely to use these devices to treat malaria conditions than those without mobile wireless devices.

Moreover, the chi-square test results also show a significant association between mHealth intervention availability and its use to manage malaria conditions (p< 0.05). Healthcare professionals with mHealth interventions were more likely to use such interventions to support malaria management than those without mHealth. The results further illustrate a significant relationship between short message services (SMS) and their use to manage hypertension cases (p< 0.05). Healthcare workers who stipulated that they have SMS applications were more likely to use such intervention to manage hypertension conditions than those without SMS services. Also, the chi-square test results suggest a significant relationship between mobile apps and their use to manage TB (p< 0.05), diabetes (p< 0.05), and disease diagnosis (p< 0.05). Healthcare professionals who indicated that they have mobile apps were likely to use them for diagnosing or screening diseases and managing TB and diabetes conditions than others with no mobile apps. The chi-square test results demonstrate a significant association between toll-free lines and their usage for managing TB (p< 0.05) and HIV (p< 0.05) conditions. Healthcare workers who suggested that they have toll-free lines were likely to use this intervention to support the treatment of TB and HIV conditions than others without toll-free lines.

### Association between health infrastructure availability or healthcare workforce competency and usefulness of mHealth applications

A cross-sectional tabulation was performed between healthcare infrastructure’s availability or healthcare workforce competency and usefulness of mHealth applications using a chi-square test (Table S 5 supplementary material file). The chi-square test results suggest a significant relationship between mobile wireless devices’ availability and managing non-communicable diseases (NCDs) (p< 0.05), communicable diseases (p< 0.05), reminders for treatment adherence procedures (p< 0.05), clinic appointments (p< 0.05), follow-ups (p< 0.05) and treating and managing diseases (p< 0.05) to support healthcare provision. Healthcare workers with mobile wireless devices were likely to use these devices to manage communicable and non-communicable diseases, reminders for treatment adherence procedures, clinic appointments, and follow-ups compared with those without mobile wireless devices.

The chi-square test results also found a significant association between the availability of mHealth intervention and its use as reminders for treatment adherence procedures (p< 0.05), clinic appointments (p< 0.05), follow-ups (p< 0.05), and test result notifications (p< 0.05) to promote healthcare delivery. Healthcare professionals who indicated that they mHealth interventions were likely to use these interventions as reminders for treatment adherence procedures, clinic appointments, follow-ups, and test result notifications than others with no mHealth interventions. The results further illustrate a significant relationship between short message services (SMS) and their use to manage NCDs (p< 0.05) and follow-ups to promote treatment compliance (p< 0.05). Healthcare workers with SMS interventions were more likely to use such interventions to manage NCDs and follow-ups than those without mHealth.

### Association between health infrastructure availability or healthcare workforce competency and ease of use of mHealth applications

A cross-sectional tabulation was done between the availability of healthcare infrastructure or healthcare workforce competency and ease of use of mHealth applications using a chi-square test (Table S 6 supplementary material file). The chi-square test results reveal a significant relationship between mobile wireless devices’ availability and its ease of using mHealth for disease diagnosis (p< 0.05), treatment support (p< 0.05), and its flexibility (p< 0.05) to support healthcare services. Healthcare workers who indicated that they have mobile wireless devices were more likely to find it easier and flexible to use them for disease diagnosis and treatment support than those without mobile wireless devices. In addition to that, the chi-square test results show a significant association between the availability of mHealth intervention and its ease of using mHealth for treatment support (p< 0.05) and its flexibility (p< 0.05) to promote healthcare delivery. Healthcare professionals with mHealth interventions were likely to find it easier and flexible to use these mHealth interventions to support patients’ disease diagnosis and treatment conditions than others with no mHealth interventions.

The results further found a significant relationship between short message services (SMS) and its ease of using mHealth for treatment support (p< 0.05) and its flexibility (p< 0.05) to enhance the provision of quality healthcare. Healthcare professionals with SMS interventions were more likely to find it easier and flexible to use such mHealth interventions for disease diagnosis and treatment support than those without mHealth. Also, the chi-square test results show a significant relationship between phone calls and its ease of using mHealth for disease diagnosis (p< 0.05), treatment support (p< 0.05), flexibility (p< 0.05), becoming skillful in using mHealth (p< 0.05) and easy to learn how to use mHealth (p< 0.05). Healthcare workers who indicated that they use phone call intervention were likely to find it easier and flexible to use such applications for disease diagnosis and treatment support than other healthcare workers without access to voice calls.

### Association between health infrastructure availability or healthcare workforce competency and user satisfaction of mHealth

A cross-sectional tabulation was done between healthcare infrastructure’s availability or healthcare workforce competency and user satisfaction of mHealth applications using a chi-square test (Table S 7 supplementary material file). The chi-square test results show a significant relationship between mobile wireless devices’ availability and confidence (p< 0.05) and completely satisfied (p< 0.05) using mHealth for disease diagnosis and treatment support. Healthcare professionals who were confident and completely satisfied with mHealth were more likely to use these mobile wireless devices for disease diagnosis and treatment support than those with no confidence in mHealth. Also, the chi-square test results illustrate a significant association between the availability of mHealth intervention and comfortable using mHealth (p< 0.05), confidence in mHealth (p< 0.05), and increase quality healthcare (p< 0.05). Healthcare professionals with mHealth interventions who were comfortable and confident in mHealth were likely to use such applications to boost quality healthcare delivery than others with no mHealth interventions.

The results further found a significant relationship between short message services (SMS) and completely satisfied with mHealth applications (p< 0.05). Healthcare workers with SMS applications were more likely to be happy with mHealth interventions for disease diagnosis and treatment support than others with no SMS application access. In addition to that, the chi-square test results indicate a significant association between phone calls and their comfortability (p< 0.05) and increase quality healthcare (p< 0.05). Healthcare professionals who suggested that they use phone call intervention were likely to be comfortable in using mHealth to boost quality healthcare delivery than those without access to voice calls. Again, the chi-square test results illustrate a significant relationship between mobile apps and completely satisfied in using mHealth applications (p< 0.05). Healthcare workers with mobile apps were more likely to be happy with mHealth for disease diagnosis and treatment support than others with no mobile apps. The chi-square test results reveal a significant association between toll-free intervention and its comfortability using mHealth (p< 0.05). Healthcare professionals who have access to toll-free lines were more likely to be comfortable using mHealth than those without toll-free lines.

### Association between health infrastructure availability or healthcare workforce competency and behavioural intention to use mHealth

A cross-sectional tabulation was performed between the availability of healthcare infrastructure or healthcare workforce competency and behavioural intention to use mHealth using a chi-square test (Table S 7 supplementary material file). The chi-square test results illustrate a significant relationship between mobile wireless devices’ availability and always use mHealth for disease diagnosis and treatment support (p< 0.05). Healthcare professionals with mobile wireless devices were more likely to use mHealth for disease diagnosis and treatment support than others with no mobile wireless devices. Additionally, the chi-square test results found a significant association between mHealth intervention availability and always use mHealth for disease diagnosis and treatment support (p< 0.05). Healthcare workers with mHealth interventions were likely to use mHealth for disease diagnosis than others with no mHealth interventions.

Furthermore, the results also found a significant relationship between SMS and the ability to use mHealth to treat and manage patients’ conditions (p< 0.05). Healthcare professionals with SMS interventions were more likely to use mHealth to treat and manage patients’ needs than others with no SMS intervention access. The chi-square test results found a significant association between phone calls and intention to use mHealth for disease diagnosis and treatment support (p< 0.05). Healthcare workers who indicated that they use phone call intervention intended to use mHealth for disease diagnosis and treatment support than those with no access to voice calls. Again, the chi-square test results demonstrate a significant relationship between mobile apps and always use mHealth for disease diagnosis and treatment support (p< 0.05). Healthcare professionals with mobile apps were more likely to use mHealth for disease diagnosis and treatment support than those without mobile apps.

## Discussion

This study aimed to examine the availability and use of mHealth applications or technologies for disease diagnosis and treatment support by healthcare workers in Ghana. In this study, 62.81% of healthcare professionals indicated that mHealth applications are available to them, while 37.19% do not have access to mHealth applications. In this study, 98.38% of healthcare professionals are currently using mHealth applications to support healthcare delivery. In this current study, the findings showed that mobile wireless devices such as simple mobile phones, smartphones, and tablets are readily available to healthcare professionals in Ghana’s Ashanti region. The results also revealed that mHealth applications such as phone or voice calls, SMS, mobile apps, and toll-free lines are available to healthcare workers in this region and are currently being used to support healthcare delivery. The study results further illustrated that healthcare professionals extensively use mHealth applications to screen or diagnose existing many disease conditions in this region.

Additionally, this study’s results demonstrated that healthcare workers in this part of Ghana currently use mHealth applications to treat HIV, TB, hypertension, diabetes, malaria, and diarrhoea conditions. Also, the study results revealed that healthcare professionals continuously use mHealth applications to support the provision of healthcare due to the constant supply of power. Moreover, the study findings suggested that most healthcare professionals have the requisite skills and competence in using mHealth applications for diagnostics and treatment procedures of disease conditions. Furthermore, the study results demonstrated a low-level use of mHealth applications for disease diagnosis and treatment support by healthcare professionals at the rural primary healthcare clinics.

A study conducted in the USA largely agrees with this current survey study where healthcare workers use mHealth applications to treat and manage chronic diseases such as HIV, TB, cancer, hypertension, diabetes, and others (48). This current survey results fully support the findings from similar surveys conducted in primary care clinics in the USA (50, 51). In their studies, most healthcare workers were comfortable and confident in using mHealth applications to support their patients’ healthcare needs (50, 51). The study findings demonstrate that healthcare workers use mHealth applications to promote medication adherence, clinic appointments, and follow-ups. This corroborates with the study findings from a similar study conducted by Belcher et al. in Saudi Arabia, where mHealth applications improved the treatment of diabetes, clinic appointments, and check-ups (52).

This current study’s limitations may include respondents’ inclusion was based on patient consent, which may have introduced selection bias into the study sample. Due to the limited funding for the data collection, only 285 participants were enrolled from the numerous primary healthcare clinics in this region. Our current study results may not be generalized beyond the Ashanti region of Ghana among healthcare professionals using mHealth applications for disease diagnosis and treatment support. Despite all these limitations, our current study is the first comprehensive research to the best of our knowledge on the availability and use of mHealth applications for disease diagnosis and treatment support by healthcare professionals in the Ashanti region of Ghana. The study helped determine the current availability and use of mHealth applications by healthcare professionals for diagnostics and treatment procedures of diseases in the Ashanti region. This could guide policymakers in formulating guidelines on the utilization mHealth technologies to promote quality healthcare delivery.

This current study achieved its primary objective and demonstrated a gap in mHealth for disease diagnosis and treatment support at the rural primary healthcare clinics in Ghana’s Ashanti region. This means that policymakers and implementors should adopt various strategies to facilitate the implementation of mHealth applications for disease diagnosis and treatment support in such resource-constrained settings and enhance their scale-ups. Given this, we recommend a proposed framework for improving the implementation of mHealth for disease diagnosis and treatment support in low-and middle-income countries (LMICs) (53). The results showed that mHealth applications are generally available to healthcare professionals and are being utilized for disease diagnosis and treatment support of patients’ conditions. This is a good sign that the continuous use of mHealth should be strengthened to promote quality healthcare delivery as recommended by the World Health Organization (WHO) 2019 guidelines on digital health (54).

The study results demonstrated a low-level use of mHealth applications for disease diagnosis and treatment support by healthcare professionals at the rural primary healthcare clinics. To this end, we encourage policymakers to deliberately implement mHealth at rural primary health clinics to support disease diagnosis and treatment procedures of patients’ conditions. The study findings showed that healthcare professionals employed mHealth applications to treat some common diseases such as HIV, TB, hypertension, and diabetes. We recommend that more primary studies be conducted focused on using mHealth interventions to treat and manage other diseases such as cancer, stroke, chronic respiratory conditions, asthma, and others in this region. Moreover, the study findings indicated that most healthcare professionals use mHealth applications to screen or diagnose several common disease conditions in this region. Hence, we encouraged healthcare professionals to use mHealth interventions to screen or diagnose several other neglected tropical diseases to enhance early detection to initiate proper treatment processes.

## Conclusion

The study revealed that mHealth applications are primarily available to healthcare professionals to promote quality healthcare delivery in the Ashanti region. The findings showed that healthcare professionals use mHealth applications to screen or diagnose, treat, and manage several common disease conditions at primary healthcare clinics. The results of the study, in addition, demonstrated a low-level use of mHealth applications for disease diagnosis and treatment support by healthcare professionals at the rural primary healthcare clinics. Future studies are recommended to examine the availability and use of mHealth applications for disease diagnosis and treatment support by healthcare professionals at the rural primary healthcare clinics.

## Supporting information

study site

survey tool

## Data Availability

Data for this study are the property of the University of KwaZulu-Natal and can be made available publicly. All interested persons can access the data set from the author Ernest Osei via this email address: and the University of KwaZulu-Natal Biomedical Research Ethics Committee (BREC) using the following contacts: The Chairperson Biomedical Research Ethics Administration Research Office, Westville Campus, Govan Mbeki Building University of KwaZulu-Natal P/Bag X54001, Durban, 4000
KwaZulu-Natal, South Africa Tel.: +27 31260 4769 Fax: +27 31260 4609.

## Abbreviation

GHS: Ghana Health Service
GoG: Government of Ghana
HIV: Human Immunodeficiency Virus
LMICs: Low-and middle-income countries
NCDs: Non-communicable diseases
RHD: Regional Health Directorate
SMS: Short message services
SSA: Sub-Saharan African
TB: Tuberculosis
WHO: World Health Organization

## Acknowledgments

We thank the University of KwaZulu-Natal for providing us with essential research resources during this study. The authors are also grateful to all the staff members at the primary healthcare facilities who participated in this study. The authors would like to thank the Authorities of the Ashanti Regional Health Directorate, the District Health Management Teams, and all the PHCs managers for permitting us to conduct this study. Finally, we are grateful to the Department of Public Health Medicine staff for their support in diverse ways.

## Contributors

EO and TPM-T conceptualized the study and developed the analytical strategy. EO collected the data, processed the data, performed the statistical analysis, interpreted the results, and wrote the first draft of the study. KA and BT contributed to the statistical analysis and interpretation of the results. TPM-T contributed to the analytical strategy, to the interpretation of the results, and did critical revisions. All authors contributed to the writing of the manuscript and approved the final version.

## Funding

This study did not receive any internal or external funding.

## Competing interests

None declared.

## Patient and Public Involvement

Patients and/or the public were involved in the design, or conduct, or reporting, or dissemination plans of this research.

## Patient consent for publication

Not required.

## Data Availability

Data for this study are the property of the University of KwaZulu-Natal and can be made available publicly. All interested persons can access the data set from the author Ernest Osei via this email address: and the University of KwaZulu-Natal Biomedical Research Ethics Committee (BREC) using the following contacts: The Chairperson Biomedical Research Ethics Administration Research Office, Westville Campus, Govan Mbeki Building University of KwaZulu-Natal P/Bag X54001, Durban, 4000 KwaZulu-Natal, South Africa Tel.: +27 31260 4769 Fax: +27 31260 4609.

## Supplementary files

**Additional file 1:** Distribution of primary healthcare facilities sampled in the Ashanti Region

**Additional file 2:** Survey tool

**Table S 1:** Characteristics of participants from the 100 healthcare facilities surveyed in Ashanti region

**Table S 2:** Availability of mobile health for diagnostics and treatment support in the Ashanti region

**Table S 3:** Use of mobile health for diagnostics and treatment support in the Ashanti region

**Table S 4:** Chi-Square Tests Results of the relationship between the available health infrastructure or healthcare workforce competency and ownership of mobile wireless devices

**Table S 5:** Chi-Square Tests Results of the relationship between the available health infrastructure or healthcare workforce competency and usefulness of mHealth applications

**Table S 6:** Chi-Square Tests Results of the relationship between the available health infrastructure or healthcare workforce competency and ease of use of mHealth applications

**Table S 7:** Chi-Square Tests Results of the relationship between the available health infrastructure or healthcare workforce competency and user satisfaction and behavioural intention to use mHealth

**Supplementary figure 1:** Odds ratio showing the association on the availability of mobile apps, toll-free, supply of power, support systems and others for disease diagnosis and treatment support by health workers in Ashanti region, Ghana.

**Supplementary figure 2:** Odds ratio showing the association on the use of mHealth applications for the management and treatment of hypertension, diabetes, cancer, malaria, monitor patients’ conditions and others by health workers in Ashanti region, Ghana.

## Notes

### Competing Interest Statement

The authors have declared no competing interest.

### Author Declarations

Biomedical Research Ethics Committee, University of KwaZulu-Natal (approval reference no. BREC/00000202/2019), Ghana Health Service Ethics Review Committee (approval reference no. GHS-ERC006/11/19

