## supplementary figure 1 for "Availability and Use of Mobile Health Technology for Disease Diagnosis and Treatment Support by Health Workers in the Ashanti Region of Ghana: A Cross-sectional Survey"

Odds ratio

|  | variables |
| --- | --- |
| 31-40 years | 1 |
| Above 40 years | 2 |
| male | 3 |
| female | 4 |
| doctors | 5 |
| nurses | 6 |
| pharmacist | 7 |
| laboratory scientists | 8 |
| district hospital | 9 |
| health centre | 10 |
| clinic | 11 |
| Number of health professionals | 12 |
| Number of patients per week | 13 |
|  | 14 |

|  | odds |
| --- | --- |
| 1.46 | 1 |
| 2.21 | 2 |
| 1 | 3 |
| 1.27 | 4 |
| 1 | 5 |
| 1.16 | 6 |
| 0.84 | 7 |
| 0.83 | 8 |
| 1 | 9 |
| 0 | 10 |
| 0 | 11 |
| 1 | 12 |
| 1.16 | 13 |
| 1 | 14 |

Odds ratio

|  | variables |
| --- | --- |
| 31-40 years | 1 |
| Above 40 years | 2 |
| male | 3 |
| female | 4 |
| doctors | 5 |
| nurses | 6 |
| pharmacist | 7 |
| laboratory scientists | 8 |
| district hospital | 9 |
| health centre | 10 |
| clinic | 11 |
| Number of health professionals | 12 |
| Number of patients per week | 13 |
|  | 14 |

|  | odds |
| --- | --- |
| 17.8 | 1 |
| 17.6 | 2 |
| 1 | 3 |
| 1.02 | 4 |
| 1 | 5 |
| 1.48 | 6 |
| 0.96 | 7 |
| 0.84 | 8 |
| 1 | 9 |
| 23.8 | 10 |
| 8.47 | 11 |
| 1 | 12 |
| 1.01 | 13 |
| 0.97 | 14 |

Odds ratio

|  | variables |
| --- | --- |
| 31-40 years | 1 |
| Above 40 years | 2 |
| male | 3 |
| female | 4 |
| doctors | 5 |
| nurses | 6 |
| pharmacist | 7 |
| laboratory scientists | 8 |
| district hospital | 9 |
| health centre | 10 |
| clinic | 11 |
| Number of health professionals | 12 |
| Number of patients per week | 13 |
|  | 14 |

|  | odds |
| --- | --- |
| 0.66 | 1 |
| 1.49 | 2 |
| 1 | 3 |
| 0.81 | 4 |
| 1 | 5 |
| 1 | 6 |
| 1.52 | 7 |
| 1.27 | 8 |
| 1 | 9 |
| 2.16 | 10 |
| 0.59 | 11 |
| 1 | 12 |
| 1.01 | 13 |
| 0.99 | 14 |

Odds ratio

|  | variables |
| --- | --- |
| 31-40 years | 1 |
| Above 40 years | 2 |
| male | 3 |
| female | 4 |
| doctors | 5 |
| nurses | 6 |
| pharmacist | 7 |
| laboratory scientists | 8 |
| district hospital | 9 |
| health centre | 10 |
| clinic | 11 |
| Number of health professionals | 12 |
| Number of patients per week | 13 |
|  | 14 |

|  | odds |
| --- | --- |
| 0.7229 | 1 |
| 0.823 | 2 |
| 1 | 3 |
| 0.917 | 4 |
| 1 | 5 |
| 1.132 | 6 |
| 1.236 | 7 |
| 0.924 | 8 |
| 1 | 9 |
| 0.872 | 10 |
| 0.133 | 11 |
| 1 | 12 |
| 1.073 | 13 |
| 0.9778 | 14 |

Odds ratio

|  | variables |
| --- | --- |
| 31-40 years | 1 |
| Above 40 years | 2 |
| male | 3 |
| female | 4 |
| doctors | 5 |
| nurses | 6 |
| pharmacist | 7 |
| laboratory scientists | 8 |
| district hospital | 9 |
| health centre | 10 |
| clinic | 11 |
| Number of health professionals | 12 |
| Number of patients per week | 13 |
|  | 14 |

|  | odds |
| --- | --- |
| 1.418 | 1 |
| 1.769 | 2 |
| 1 | 3 |
| 0.985 | 4 |
| 1 | 5 |
| 1.242 | 6 |
| 1.69 | 7 |
| 1.28 | 8 |
| 1 | 9 |
| 0.119 | 10 |
| 0.409 | 11 |
| 1 | 12 |
| 1.057 | 13 |
| 1 | 14 |

Odds ratio

|  | variables |
| --- | --- |
| 31-40 years | 1 |
| Above 40 years | 2 |
| male | 3 |
| female | 4 |
| doctors | 5 |
| nurses | 6 |
| pharmacist | 7 |
| laboratory scientists | 8 |
| district hospital | 9 |
| health centre | 10 |
| clinic | 11 |
| Number of health professionals | 12 |
| Number of patients per week | 13 |
|  | 14 |

|  | odds |
| --- | --- |
| 0.978 | 1 |
| 1.491 | 2 |
| 1 | 3 |
| 1.075 | 4 |
| 1 | 5 |
| 0.386 | 6 |
| 0.345 | 7 |
| 0.487 | 8 |
| 1 | 9 |
| 29.87 | 10 |
| 13.53 | 11 |
| 1 | 12 |
| 0.907 | 13 |
| 0.997 | 14 |

Odds ratio

|  | variables |
| --- | --- |
| 31-40 years | 1 |
| Above 40 years | 2 |
| male | 3 |
| female | 4 |
| doctors | 5 |
| nurses | 6 |
| pharmacist | 7 |
| laboratory scientists | 8 |
| district hospital | 9 |
| health centre | 10 |
| clinic | 11 |
| Number of health professionals | 12 |
| Number of patients per week | 13 |
|  | 14 |

|  | odds |
| --- | --- |
| 0.298 | 1 |
| 0.604 | 2 |
| 1 | 3 |
| 1.208 | 4 |
| 1 | 5 |
| 1.065 | 6 |
| 0.723 | 7 |
| 1.243 | 8 |
| 1 | 9 |
| 21.66 | 10 |
| 2.777 | 11 |
| 1 | 12 |
| 0.969 | 13 |
| 0.995 | 14 |

Odds ratio

|  | variables |
| --- | --- |
| 31-40 years | 1 |
| Above 40 years | 2 |
| male | 3 |
| female | 4 |
| doctors | 5 |
| nurses | 6 |
| pharmacist | 7 |
| laboratory scientists | 8 |
| district hospital | 9 |
| health centre | 10 |
| clinic | 11 |
| Number of health professionals | 12 |
| Number of patients per week | 13 |
|  | 14 |

|  | odds |
| --- | --- |
| 0.595 | 1 |
| 1.805 | 2 |
| 1 | 3 |
| 0.671 | 4 |
| 1 | 5 |
| 1.153 | 6 |
| 0.345 | 7 |
| 0.654 | 8 |
| 1 | 9 |
| 0.623 | 10 |
| 0.63 | 11 |
| 1 | 12 |
| 1.196 | 13 |
| 1.019 | 14 |

Odds ratio

|  | variables |
| --- | --- |
| 31-40 years | 1 |
| Above 40 years | 2 |
| male | 3 |
| female | 4 |
| doctors | 5 |
| nurses | 6 |
| pharmacist | 7 |
| laboratory scientists | 8 |
| district hospital | 9 |
| health centre | 10 |
| clinic | 11 |
| Number of health professionals | 12 |
| Number of patients per week | 13 |
|  | 14 |

|  | odds |
| --- | --- |
| 0.654 | 1 |
| 0.871 | 2 |
| 1 | 3 |
| 1.209 | 4 |
| 1 | 5 |
| 1.143 | 6 |
| 0.787 | 7 |
| 0.792 | 8 |
| 1 | 9 |
| 24.3 | 10 |
| 10.86 | 11 |
| 1 | 12 |
| 0.948 | 13 |
| 1.002 | 14 |

**Supplementary figure 1:** Odds ratio showing the association on the availability of mobile apps, toll-free, supply of power, support systems and others for disease diagnosis and treatment support by health workers in Ashanti region, Ghana.
