## supplementary figure 2 for "Availability and Use of Mobile Health Technology for Disease Diagnosis and Treatment Support by Health Workers in the Ashanti Region of Ghana: A Cross-sectional Survey"

Odds ratio

|  | variables |
| --- | --- |
| 31-40 years | 1 |
| Above 40 years | 2 |
| male | 3 |
| female | 4 |
| doctors | 5 |
| nurses | 6 |
| pharmacist | 7 |
| laboratory scientists | 8 |
| district hospital | 9 |
| health centre | 10 |
| clinic | 11 |
| Number of health professionals | 12 |
| Number of patients per week | 13 |
|  | 14 |

|  | odds |
| --- | --- |
| 2.28 | 1 |
| 2.12 | 2 |
| 1 | 3 |
| 0.84 | 4 |
| 1 | 5 |
| 0.78 | 6 |
| 0.53 | 7 |
| 1.19 | 8 |
| 1 | 9 |
| 2.47 | 10 |
| 1.06 | 11 |
| 1 | 12 |
| 1 | 13 |
| 0.99 | 14 |

Odds ratio

|  | variables |
| --- | --- |
| 31-40 years | 1 |
| Above 40 years | 2 |
| male | 3 |
| female | 4 |
| doctors | 5 |
| nurses | 6 |
| pharmacist | 7 |
| laboratory scientists | 8 |
| district hospital | 9 |
| health centre | 10 |
| clinic | 11 |
| Number of health professionals | 12 |
| Number of patients per week | 13 |
|  | 14 |

|  | odds |
| --- | --- |
| 3.75 | 1 |
| 5.75 | 2 |
| 1 | 3 |
| 1.37 | 4 |
| 1 | 5 |
| 0.81 | 6 |
| 0.57 | 7 |
| 1.26 | 8 |
| 1 | 9 |
| 0.12 | 10 |
| 0.45 | 11 |
| 1 | 12 |
| 1.04 | 13 |
| 0.99 | 14 |

Odds ratio

|  | variables |
| --- | --- |
| 31-40 years | 1 |
| Above 40 years | 2 |
| male | 3 |
| female | 4 |
| doctors | 5 |
| nurses | 6 |
| pharmacist | 7 |
| laboratory scientists | 8 |
| district hospital | 9 |
| health centre | 10 |
| clinic | 11 |
| Number of health professionals | 12 |
| Number of patients per week | 13 |
|  | 14 |

|  | odds |
| --- | --- |
| 0.16 | 1 |
| 0.26 | 2 |
| 1 | 3 |
| 1.94 | 4 |
| 1 | 5 |
| 1.66 | 6 |
| 1.28 | 7 |
| 1.13 | 8 |
| 1 | 9 |
| 18.1 | 10 |
| 3.89 | 11 |
| 1 | 12 |
| 0.98 | 13 |
| 1.2 | 14 |

Odds ratio

|  | variables |
| --- | --- |
| 31-40 years | 1 |
| Above 40 years | 2 |
| male | 3 |
| female | 4 |
| doctors | 5 |
| nurses | 6 |
| pharmacist | 7 |
| laboratory scientists | 8 |
| district hospital | 9 |
| health centre | 10 |
| clinic | 11 |
| Number of health professionals | 12 |
| Number of patients per week | 13 |
|  | 14 |

|  | odds |
| --- | --- |
| 6.05 | 1 |
| 11.1 | 2 |
| 1 | 3 |
| 0.04 | 4 |
| 1 | 5 |
| 0.85 | 6 |
| 0.14 | 7 |
| 0.85 | 8 |
| 1 | 9 |
| 0 | 10 |
| 0 | 11 |
| 1 | 12 |
| 0.96 | 13 |
| 0.97 | 14 |

Odds ratio

|  | variables |
| --- | --- |
| 31-40 years | 1 |
| Above 40 years | 2 |
| male | 3 |
| female | 4 |
| doctors | 5 |
| nurses | 6 |
| pharmacist | 7 |
| laboratory scientists | 8 |
| district hospital | 9 |
| health centre | 10 |
| clinic | 11 |
| Number of health professionals | 12 |
| Number of patients per week | 13 |
|  | 14 |

|  | odds |
| --- | --- |
| 1.04 | 1 |
| 1.24 | 2 |
| 1 | 3 |
| 0.96 | 4 |
| 1 | 5 |
| 0.99 | 6 |
| 0.61 | 7 |
| 1.41 | 8 |
| 1 | 9 |
| 0.36 | 10 |
| 0.33 | 11 |
| 1 | 12 |
| 1 | 13 |
| 0.99 | 14 |

Odds ratio

|  | variables |
| --- | --- |
| 31-40 years | 1 |
| Above 40 years | 2 |
| male | 3 |
| female | 4 |
| doctors | 5 |
| nurses | 6 |
| pharmacist | 7 |
| laboratory scientists | 8 |
| district hospital | 9 |
| health centre | 10 |
| clinic | 11 |
| Number of health professionals | 12 |
| Number of patients per week | 13 |
|  | 14 |

|  | odds |
| --- | --- |
| 0.67 | 1 |
| 0.79 | 2 |
| 1 | 3 |
| 1.32 | 4 |
| 1 | 5 |
| 1.19 | 6 |
| 1.11 | 7 |
| 2.29 | 8 |
| 1 | 9 |
| 0.35 | 10 |
| 0.42 | 11 |
| 1 | 12 |
| 0.99 | 13 |
| 1 | 14 |

|  | variables |
| --- | --- |
| 31-40 years | 1 |
| Above 40 years | 2 |
| male | 3 |
| female | 4 |
| doctors | 5 |
| nurses | 6 |
| pharmacist | 7 |
| laboratory scientists | 8 |
| district hospital | 9 |
| health centre | 10 |
| clinic | 11 |
| Number of health professionals | 12 |
| Number of patients per week | 13 |
|  | 14 |

|  | odds |
| --- | --- |
| 1.06 | 1 |
| 7.25 | 2 |
| 1 | 3 |
| 1.48 | 4 |
| 1 | 5 |
| 0.93 | 6 |
| 0.45 | 7 |
| 1.32 | 8 |
| 1 | 9 |
| 1.7 | 10 |
| 3.96 | 11 |
| 1 | 12 |
| 1 | 13 |
| 0.99 | 14 |

Odds ratio

|  | variables |
| --- | --- |
| 31-40 years | 1 |
| Above 40 years | 2 |
| male | 3 |
| female | 4 |
| doctors | 5 |
| nurses | 6 |
| pharmacist | 7 |
| laboratory scientists | 8 |
| district hospital | 9 |
| health centre | 10 |
| clinic | 11 |
| Number of health professionals | 12 |
| Number of patients per week | 13 |
|  | 14 |

|  | odds |
| --- | --- |
| 0.74 | 1 |
| 0.92 | 2 |
| 1 | 3 |
| 1.73 | 4 |
| 1 | 5 |
| 0.87 | 6 |
| 1 | 7 |
| 1.69 | 8 |
| 1 | 9 |
| 0.28 | 10 |
| 0.54 | 11 |
| 1 | 12 |
| 1.01 | 13 |
| 1.01 | 14 |

Odds ratio

|  | variables |
| --- | --- |
| 31-40 years | 1 |
| Above 40 years | 2 |
| male | 3 |
| female | 4 |
| doctors | 5 |
| nurses | 6 |
| pharmacist | 7 |
| laboratory scientists | 8 |
| district hospital | 9 |
| health centre | 10 |
| clinic | 11 |
| Number of health professionals | 12 |
| Number of patients per week | 13 |
|  | 14 |

|  | odds |
| --- | --- |
| 0.64 | 1 |
| 1.04 | 2 |
| 1 | 3 |
| 1.22 | 4 |
| 1 | 5 |
| 1.35 | 6 |
| 1.17 | 7 |
| 0.87 | 8 |
| 1 | 9 |
| 1.6 | 10 |
| 1.41 | 11 |
| 1 | 12 |
| 1 | 13 |
| 1 | 14 |

|  | variables |
| --- | --- |
| 31-40 years | 1 |
| Above 40 years | 2 |
| male | 3 |
| female | 4 |
| doctors | 5 |
| nurses | 6 |
| pharmacist | 7 |
| laboratory scientists | 8 |
| district hospital | 9 |
| health centre | 10 |
| clinic | 11 |
| Number of health professionals | 12 |
| Number of patients per week | 13 |
|  | 14 |

|  | odds |
| --- | --- |
| 3.05 | 1 |
| 3.64 | 2 |
| 1 | 3 |
| 1.17 | 4 |
| 1 | 5 |
| 0.56 | 6 |
| 1.48 | 7 |
| 0.64 | 8 |
| 1 | 9 |
| 6.43 | 10 |
| 1.34 | 11 |
| 1 | 12 |
| 0.98 | 13 |
| 0.99 | 14 |

**Supplementary figure 2:** Odds ratio showing the association on the use of mHealth applications for the management and treatment of hypertension, diabetes, cancer, malaria, monitor patients’ conditions and others by health workers in Ashanti region, Ghana.

Odds ratio

|  | variables |
| --- | --- |
| 31-40 years | 1 |
| Above 40 years | 2 |
| male | 3 |
| female | 4 |
| doctors | 5 |
| nurses | 6 |
| pharmacist | 7 |
| laboratory scientists | 8 |
| district hospital | 9 |
| health centre | 10 |
| clinic | 11 |
| Number of health professionals | 12 |
| Number of patients per week | 13 |
|  | 14 |

|  | odds |
| --- | --- |
| 2.16 | 1 |
| 2.86 | 2 |
| 1 | 3 |
| 0.66 | 4 |
| 1 | 5 |
| 0.38 | 6 |
| 0.79 | 7 |
| 0.69 | 8 |
| 1 | 9 |
| 0.88 | 10 |
| 1.5 | 11 |
| 1 | 12 |
| 1.01 | 13 |
| 2.99 | 14 |

Odds ratio

|  | variables |
| --- | --- |
| 31-40 years | 1 |
| Above 40 years | 2 |
| male | 3 |
| female | 4 |
| doctors | 5 |
| nurses | 6 |
| pharmacist | 7 |
| laboratory scientists | 8 |
| district hospital | 9 |
| health centre | 10 |
| clinic | 11 |
| Number of health professionals | 12 |
| Number of patients per week | 13 |
|  | 14 |

|  | odds |
| --- | --- |
| 0.71 | 1 |
| 0.91 | 2 |
| 1 | 3 |
| 2.33 | 4 |
| 1 | 5 |
| 1.24 | 6 |
| 1.39 | 7 |
| 2.07 | 8 |
| 1 | 9 |
| 0.76 | 10 |
| 0.39 | 11 |
| 1 | 12 |
| 0.98 | 13 |
| 1.01 | 14 |
