## supplementary table 1 for "Availability and Use of Mobile Health Technology for Disease Diagnosis and Treatment Support by Health Workers in the Ashanti Region of Ghana: A Cross-sectional Survey"

**Supplementary information**

**Table S 1:** Characteristics of participants from the 100 healthcare facilities surveyed in Ashanti region

| Variable | Frequency (N = 285) | Percentage (%) |
| --- | --- | --- |
| Age |  |  |
| (20-30) | 120 | 42.11 |
| (31-40) | 137 | 48.07 |
| (41-50) | 27 | 9.47 |
| (51-60) | 1 | 0.35 |
| Sex |  |  |
| Male | 146 | 51.23 |
| Female | 139 | 48.77 |
| Categories of health professionals |  |  |
| Medical doctor | 44 | 15.44 |
| Physician assistant | 65 | 22.81 |
| Midwife | 7 | 2.46 |
| General nurse | 82 | 28.7 |
| Community health nurse | 26 | 9.12 |
| Laboratory scientist/technicians | 31 | 10.88 |
| Pharmacist/dispensing technicians | 30 | 10.53 |
|  | **Mean (standard deviation)** | **95% Confidence Interval** |
| Total number of health professionals | 57.8 ± 30.1 | 20-98 |
| Number of patients per week | 175.4 ± 68.4 | 74-372 |

**Source: Author’s computation based on data obtained from the field survey, 2020**
