## supplementary table 2 for "Availability and Use of Mobile Health Technology for Disease Diagnosis and Treatment Support by Health Workers in the Ashanti Region of Ghana: A Cross-sectional Survey"

| Variable | Frequency (N = 285) | | Percentage (%) |
| --- | --- | --- | --- |
| Available healthcare infrastructure | | | |
| Mobile wireless devices | |  |  |
| Yes | | 276 | 96.84 |
| No | | 9 | 3.16 |
| Mobile intervention availability | |  |  |
| Yes | | 179 | 62.81 |
| No | | 106 | 37.19 |
| Types of mobile health intervention | |  |  |
| Short Message Service | |  |  |
| Yes | | 149 | 80.54 |
| No | | 36 | 19.46 |
| Phone calls | |  |  |
| Yes | | 183 | 98.92 |
| No | | 2 | 1.08 |
| Mobile apps | |  |  |
| Yes | | 43 | 23.24 |
| No | | 142 | 76.76 |
| Multimedia Service | |  |  |
| Yes | | 2 | 1.08 |
| No | | 183 | 98.92 |
| Video conferencing | |  |  |
| Yes | | - | - |
| No | | 185 | 100 |
| Toll free lines | |  |  |
| Yes | | 37 | 20.00 |
| No | | 148 | 80.00 |
| Types of wireless mobile devices | |  |  |
| Mobile phones | |  |  |
| Yes | | 185 | 100.00 |
| No | | - | - |
| Smartphones | |  |  |
| Yes | | 133 | 71.89 |
| No | | 52 | 28.11 |
| Tablets | |  |  |
| Yes | | 107 | 57.84 |
| No | | 78 | 42.16 |
| Personal digital assistants | |  |  |
| Yes | | - | - |
| No | | 185 | 100.00 |
| Handheld devices | |  |  |
| Yes | | - | - |
| No | | 185 | 100.00 |
| Patient monitoring devices | |  |  |
| Yes | | 2 | 1.08 |
| No | | 183 | 98.92 |
| Watches | |  |  |
| Yes | | - | - |
| No | | 185 | 100.00 |
| Continuous supply of power | |  |  |
| Yes | | 149 | 80.54 |
| No | | 36 | 19.46 |
| Available support systems | |  |  |
| Yes | | 106 | 57.30 |
| No | | 79 | 42.70 |
| Healthcare workforce competency | |  |  |
| Requisite skills for diagnostics | |  |  |
| Yes | | 132 | 71.35 |
| No | | 53 | 28.65 |
| Competence for treatment | |  |  |
| Yes | | 164 | 88.65 |
| No | | 21 | 11.35 |

**Not applicable: Frequencies that are not up to the sample size (285) are respondents without access to some mHealth apps** **Source: Author’s computation based on data obtained from the field survey,2020**
