## supplementary table 3 for "Availability and Use of Mobile Health Technology for Disease Diagnosis and Treatment Support by Health Workers in the Ashanti Region of Ghana: A Cross-sectional Survey"

| Variable | Frequency  (N = 285) | Percentage (%) |
| --- | --- | --- |
| You and Your mobile wireless device |  |  |
| Have you ever used or currently using mHealth interventions |  |  |
| Yes | 182 | 98.38 |
| No | 3 | 1.62 |
| Type of diseases mHealth has been used for |  |  |
| HIV |  |  |
| Yes | 177 | 95.68 |
| No | 8 | 4.32 |
| TB |  |  |
| Yes | 171 | 92.43 |
| No | 14 | 7.57 |
| Hypertension |  |  |
| Yes | 99 | 53.51 |
| No | 86 | 46.49 |
| Diabetes |  |  |
| Yes | 79 | 42.70 |
| No | 106 | 57.30 |
| Stroke |  |  |
| Yes | - | - |
| No | 185 | 100.00 |
| Cancer |  |  |
| Yes | 5 | 2.70 |
| No | 180 | 97.30 |
| Chronic Respiratory Disease |  |  |
| Yes | 2 | 1.08 |
| No | 183 | 98.92 |
| Malaria |  |  |
| Yes | 93 | 50.54 |
| No | 91 | 49.46 |
| Diarrhoea |  |  |
| Yes | 17 | 9.19 |
| No | 168 | 90.81 |
| Have you ever used smart mobile wireless device |  |  |
| Health/medical information |  |  |
| Yes | 117 | 63.24 |
| No | 68 | 36.76 |
| Disease diagnosis |  |  |
| Yes | 182 | 98.38 |
| No | 3 | 1.62 |
| Treat and manage disease conditions |  |  |
| Yes | 162 | 87.57 |
| No | 23 | 12.43 |
| Treat and monitor patients' health conditions |  |  |
| Yes | 144 | 77.84 |
| No | 41 | 22.16 |
| How often are mobile wireless device used for diagnostic purposes |  |  |
| Once a month | 48 | 25.95 |
| 2 or 3 times a month | 48 | 25.95 |
| 1 to 6 times a week | 57 | 30.81 |
| Once a day or more | 32 | 17.30 |
| How often are mobile wireless device used for treatment, monitoring, and management of diseases |  |  |
| Once a month | 58 | 31.35 |
| 2 or 3 times a month | 48 | 25.95 |
| 1 to 6 times a week | 44 | 23.78 |
| Once a day or more | 35 | 18.92 |
| Usefulness of mHealth interventions |  |  |
| For monitoring patients' disease conditions |  |  |
| Yes | 248 | 87.63 |
| No | 35 | 12.37 |
| For managing non-communicable diseases |  |  |
| Yes | 189 | 66.32 |
| No | 96 | 33.68 |
| For managing communicable diseases |  |  |
| Yes | 239 | 83.86 |
| No | 46 | 16.14 |
| Reminders for treatment procedures adherence |  |  |
| Yes | 175 | 61.40 |
| No | 110 | 38.60 |
| Reminders for patients' medication adherence |  |  |
| Yes | 234 | 82.11 |
| No | 51 | 17.89 |
| Reminders to honour clinic appointments |  |  |
| Yes | 229 | 80.35 |
| No | 56 | 19.65 |
| Reminders for collection of ART and other drugs on time |  |  |
| Yes | 247 | 86.67 |
| No | 38 | 13.33 |
| For follow-ups to promote treatment compliance |  |  |
| Yes | 210 | 73.68 |
| No | 75 | 26.32 |
| For supporting patients' test result notifications |  |  |
| Yes | 187 | 65.61 |
| No | 98 | 34.39 |
| For improving treatment and management of disease conditions |  |  |
| Yes | 213 | 74.74 |
| No | 72 | 25.26 |
| For making accurate diagnostic decisions |  |  |
| Yes | 253 | 88.77 |
| No | 32 | 11.23 |
| To increase the effectiveness of treatment and management of diseases |  |  |
| Yes | 249 | 87.37 |
| No | 36 | 12.63 |
| Ease of use |  |  |
| Easy to use mHealth to support disease diagnosis |  |  |
| Yes | 262 | 91.93 |
| No | 23 | 8.07 |
| Easy to use mHealth to support the treatment of patients' disease conditions |  |  |
| Yes | 273 | 95.79 |
| No | 12 | 4.21 |
| Flexible to interact with mHealth interventions |  |  |
| Yes | 273 | 95.79 |
| No | 12 | 4.21 |
| Frustrating to interact with mHealth applications |  |  |
| Yes | 9 | 3.16 |
| No | 276 | 96.84 |
| Easy to become skillful in using mHealth |  |  |
| Yes | 267 | 93.68 |
| No | 18 | 6.32 |
| Easy to learn how to use mHealth devices for diagnosis and treatment |  |  |
| Yes | 267 | 93.68 |
| No | 18 | 6.32 |
| User satisfaction |  |  |
| Comfortable in using mHealth |  |  |
| Yes | 266 | 93.33 |
| No | 19 | 6.67 |
| Confident in using mHealth |  |  |
| Yes | 254 | 89.12 |
| No | 31 | 10.88 |
| Completely satisfied with using mHealth |  |  |
| Yes | 218 | 76.49 |
| No | 67 | 23.51 |
| Using mHealth will increase the quality of healthcare delivery |  |  |
| Yes | 268 | 94.04 |
| No | 17 | 5.96 |
| Behavioural intention to use |  |  |
| Would you use mHealth for the treatment and management of patients' disease conditions |  |  |
| Yes | 254 | 89.12 |
| No | 31 | 10.88 |
| Would you always use mHealth for disease diagnosis and treatment support |  |  |
| Yes | 224 | 78.60 |
| No | 61 | 21.40 |
| Intend to use mHealth for disease diagnosis and treatment support |  |  |
| Yes | 279 | 97.89 |
| No | 6 | 2.11 |

**Not applicable: Frequencies that are not up to the sample size (285) are respondents without access to some mHealth apps**

**Source: Author's computation based on data obtained from the field survey, 2020**
