## supplementary table 4 for "Availability and Use of Mobile Health Technology for Disease Diagnosis and Treatment Support by Health Workers in the Ashanti Region of Ghana: A Cross-sectional Survey"

**Table S 4:** Chi-Square Tests Results of the relationship between the available health infrastructure or healthcare workforce competency and ownership of mobile wireless devices

| **Availability of Health Infrastructure** | **Ownership of mobile wireless devices** | | | | | | | | | | | | | | | | | |
| --- | --- | --- | --- | --- | --- | --- | --- | --- | --- | --- | --- | --- | --- | --- | --- | --- | --- | --- |
|  | Ever used or currently using mHealth | | Malaria | | Hypertension |  | Tuberculosis (TB) | | Diabetes | | HIV | | Cancer | | Disease diagnosis | | Monitor patients' conditions | |
|  | **Chi-Square** | **p-value** | **Chi-Square** | **p-value** | **Chi-Square** | **p-value** | **Chi-Square** | **p-value** | **Chi-Square** | **p-value** | **Chi-Square** | **p-value** | **Chi-Square** | **p-value** | **Chi-Square** | **p-value** | **Chi-Square** | **p-value** |
| Availability of mobile wireless devices | 10.88 | 0.001 | 4.12 | 0.041 | - | **-** | **-** | **-** | **-** | **-** | **-** | **-** | **-** | **-** | **-** | **-** | **-** | **-** |
| mHealth intervention availability | **-** | **-** | 6.34 | 0.012 | **-** | **-** | **-** | **-** | **-** | **-** | **-** | **-** | **-** | **-** | **-** | **-** | **-** | **-** |
| SMS | - | - | **-** | **-** | 6.29 | 0.012 | **-** | **-** | **-** | **-** | **-** | **-** | **-** | **-** | **-** | **-** | **-** | **-** |
| Mobile apps | **-** | **-** | **-** | **-** | **-** | **-** | 4.59 | 0.032 | 11.5 | 0.001 |  |  | - | - | 10.1 | 0.002 | **-** | **-** |
| Toll-free | - | - | **-** | **-** | **-** | **-** | 4.95 | 0.026 | **-** | **-** | 9.44 | 0.002 | - | - | - | - |  |  |
| Smartphones | **-** | **-** | **-** | **-** | **-** | **-** | 9.81 | 0.002 | 11.4 | 0.001 | 9.09 | 0.003 | - | - | - | - | 4.65 | 0.031 |
| Supply of power | 4.34 | 0.037 | - | - | - | - | - | - | - | - | - | - | - | - | - | - | 7.25 | 0.007 |
| Support systems | - | - | - | - | - | - | 5.11 | 0.024 | - | - | - | - | 6.89 | 0.009 | 4.09 | 0.043 | - | **-** |
| Requisite skills for diagnostic purposes | - | - | - | - | - | - | 6.02 | 0.014 | - | - | 4.69 | 0.030 | - | - | - | - | - | - |
| Competence to use mHealth for treatment | - | - | - | - | - | - | 8.93 | 0.003 | - | - | - | - | 12.1 | 0.001 | - | - | - | - |

**Source: Author's computation based on data obtained from the field survey, 2020**
