## supplementary table 5 for "Availability and Use of Mobile Health Technology for Disease Diagnosis and Treatment Support by Health Workers in the Ashanti Region of Ghana: A Cross-sectional Survey"

**Table S 5:** Chi-Square Tests Results of the relationship between the available health infrastructure or healthcare workforce competency and usefulness of mHealth applications

| **Availability of Health Infrastructure** | **Usefulness of mHealth applications** | | | | | | | | | | | | | | | |
| --- | --- | --- | --- | --- | --- | --- | --- | --- | --- | --- | --- | --- | --- | --- | --- | --- |
|  | Manage non-communicable diseases | | Manage communicable diseases | | Reminders for treatment adherence procedures | | Reminders for clinic appointments | | Reminders for drugs collection | | Follow-ups to promote treatment compliance | | Treating and managing other disease conditions | | Test result notifications | |
|  | **Chi-Square** | **p-value** | **Chi-Square** | **p-value** | **Chi-Square** | **p-value** | **Chi-Square** | **p-value** | **Chi-Square** | **p-value** | **Chi-Square** | **p-value** | **Chi-Square** | **p-value** | **Chi-Square** | **p-value** |
| Availability of mobile wireless devices | 36.99 | 0.001 | 8.09 | 0.004 | 17.53 | 0.001 | 4.46 | 0.035 | 7.59 | 0.006 | 7.78 | 0.005 | 7.76 | 0.005 | - | - |
| mHealth intervention availability | - | - | - | - | 4.14 | 0.042 | 8.22 | 0.004 | - | - | 10.84 | 0.001 | - | - | 4.83 | 0.028 |
| SMS | 4.80 | 0.028 | - | - | - | - | - | - | - | - | 3.95 | 0.047 | - | - | - | - |
| Phone calls | - | - | 4.21 | 0.040 | 7.57 | 0.006 | 6.68 | 0.010 | 7.10 | 0.008 | 8.67 | 0.003 | - | - | 4.56 | 0.033 |
| Mobile apps | 6.00 | 0.014 | - | - | - | - | - | - | - | - | 10.05 | 0.002 | - | - | - | - |
| Toll-free | - | - | - | - | - | - | 5.52 | 0.019 | 9.06 | 0.003 | 14.60 | 0.001 | 7.49 | 0.006 | - | - |
| Supply of power | - | - | 6.29 | 0.012 | - | - | - | - | 8.61 | 0.003- | - | - | - | - | - | - |
| Support systems | 14.07 | 0.001 | 7.08 | 0.008 | - | - | 3.94 | 0.047 | 5.28 | 0.022 | - | - | - | - | - | - |
| Requisite skills for diagnostic purposes | 19.54 | 0.001 | 9.37 | 0.002 | - | - | - | - | - | - | - | - | - | - | - | - |
| Competence to use mHealth for treatment | 7.12 | 0.008 | 6.60 | 0.010 | - | - | - | - | - | - | - | - | - | - | - | - |

**Source: Author's computation based on data obtained from the field survey, 2020**
