## supplementary table 6 for "Availability and Use of Mobile Health Technology for Disease Diagnosis and Treatment Support by Health Workers in the Ashanti Region of Ghana: A Cross-sectional Survey"

**Table S 6:** Chi-Square Tests Results of the relationship between the available health infrastructure or healthcare workforce competency and ease of use of mHealth applications

| **Availability of Health Infrastructure** | **Ease of use of mHealth applications** | | | | | | | | | | | |
| --- | --- | --- | --- | --- | --- | --- | --- | --- | --- | --- | --- | --- |
|  | Easy to use mHealth for disease diagnosis | | Easy to use mHealth to support treatment | | Flexible to interact with mHealth applications | | Frustrating to interact with mHealth applications | | Easy to become skillful in using mHealth applications | | Easy to learn how to use mHealth devices | |
|  | **Chi-Square** | **p-value** | **Chi-Square** | **p-value** | **Chi-Square** | **p-value** | **Chi-Square** | **p-value** | **Chi-Square** | **p-value** | **Chi-Square** | **p-value** |
| Availability of mobile wireless devices | 2.78 | 0.037 | 3.82 | 0.036 | 2.41 | 0.052 | - | - | - | - | - | - |
| mHealth intervention availability | - | - | 4.20 | 0.040 | 4.47 | 0.035 | - | - | - | - | - | - |
| SMS | - | - | 4.08 | 0.043 | 5.04 | 0.025 | - | - | - | - | - | - |
| Phone calls | 11.34 | 0.001 | 17.66 | 0.001 | 31.98 | 0.001 | - | - | 11.86 | 0.001 | 24.69 | 0.001 |
| Mobile apps | - | - | - | - | - | - | - | - | 2.20 | 0.013 | - | - |
| Toll-free | - | - | - | - | 4.74 | 0.030 | - | - | - | - | - | - |
| Supply of power | - | - | - | - | - | - | - | - | 2.54 | 0.011 | - | - |
| Support systems | 11.13 | 0.001 | 3.62 | 0.050 | - | - | - | - | - | - | 2.88 | 0.049 |
| Requisite skills for diagnostic purposes | 2.80 | 0.036 | 5.96 | 0.015 | - | - | - | - | 2.73 | 0.032 | - | - |
| Competence to use mHealth for treatment | 3.33 | 0.050 | - | - | 1.49 | 0.021 | - | - | 2.06 | 0.003 | - | - |

**Source: Author's computation based on data obtained from the field survey, 2020**
