## supplementary table 7 for "Availability and Use of Mobile Health Technology for Disease Diagnosis and Treatment Support by Health Workers in the Ashanti Region of Ghana: A Cross-sectional Survey"

**Table S 7:** Chi-Square Tests Results of the relationship between the available health infrastructure or healthcare workforce competency and user satisfaction and behavioural intention to use mHealth

| **Availability of Health Infrastructure** | **User satisfaction of mHealth applications** | | | | | | | | **Behavioural intention to use mHealth applications** | | | | | |
| --- | --- | --- | --- | --- | --- | --- | --- | --- | --- | --- | --- | --- | --- | --- |
|  | Comfortable using mHealth | | Confident in using mHealth | | Completely satisfied with mHealth | | mHealth can increase quality healthcare | | Able to use mHealth to treat and manage patients' conditions | | Always use mHealth for disease diagnosis and treatment support | | Intend to use mHealth for disease diagnosis and treatment support | |
|  | **Chi-Square** | **p-value** | **Chi-Square** | **p-value** | **Chi-Square** | **p-value** | **Chi-Square** | **p-value** | **Chi-Square** | **p-value** | **Chi-Square** | **p-value** | **Chi-Square** | **p-value** |
| Availability of mobile wireless devices | - | - | 3.61 | 0.053 | 4.83 | 0.028 | - | - | - | - | 1.23 | 0.026 | - | - |
| mHealth intervention availability | 1.84 | 0.015 | 1.03 | 0.031 | - | - | 1.39 | 0.023 | - | - | 3.18 | 0.045 | - | - |
| SMS | - | - | - | - | 3.27 | 0.051 | - | - | 1.14 | 0.028 | - | - | - | - |
| Phone calls | 5.20 | 0.023 | - | - | - | - | 7.10 | 0.008 | - | - | - | - | 7.57 | 0.006 |
| Mobile apps | - | - | - | - | 2.49 | 0.011 | - | - | - | - | 1.78 | 0.018 | - | - |
| Toll-free | 2.69 | 0.040 | - | - | - | - | - | - | 1.32 | 0.025 | - | - | - | - |
| Supply of power | 2.55 | 0.011 | 2.01 | 0.015 | - | - | - | - | - | - | - | - | 1.52 | 0.046 |
| Support systems | 7.96 | 0.005 | - | - | 5.56 | 0.018 | - | - | - | - | 2.75 | 0.037 | - | - |
| Requisite skills for diagnostic purposes | 6.02 | 0.014 | - | - | - | - | 15.9 | 0.001 | 4.34 | 0.037 | - | - | - | - |
| Competence to use mHealth for treatment | 4.46 | 0.035 | - | - | 8.89 | 0.003 | - | - | - | - | 13.2 | 0.001 | - | - |

**Source: Author's computation based on data obtained from the field survey, 2020**
