## Supplementary material for "Availability and Use of Mobile Health Technology for Disease Diagnosis and Treatment Support by Health Workers in the Ashanti Region of Ghana: A Cross-sectional Survey": study site

**Additional file 1:** Distribution of primary healthcare facilities sampled in the Ashanti Region

| **Name of stratum(District) (N=43)** | **Number (N=100)** |
| --- | --- |
| Adansi Asokwa | 1 |
| Adansi Akrofroum | 2 |
| Adansi North | 2 |
| Adansi South | 2 |
| Afigya Kwabre North | 2 |
| Afigya Kwabre South | 2 |
| Ahafo Ano North | 3 |
| Ahafo Ano South-East | 2 |
| Ahafo Ano South-West | 2 |
| Amansie Central | 2 |
| Amansie South | 2 |
| Amansie West | 2 |
| Bekwai Municipal | 3 |
| Asante Akim Central | 2 |
| Asante Akim North | 2 |
| Asante Akim South | 3 |
| Mampong Municipal | 4 |
| Atwima Mponua | 2 |
| Atwima Kwawoma | 2 |
| Atwima Nwabiagya North | 3 |
| Atwima Nwabiagya South | 2 |
| Bosomtwe | 4 |
| Ejisu Municipal | 4 |
| Juaben Municipal | 2 |
| Ejura-Sekyredumase | 2 |
| Kwabre East | 3 |
| Obuasi Municipal | 2 |
| Obuasi East | 1 |
| Offinso Municipal | 2 |
| Offinso North | 2 |
| Bosome Freho | 2 |
| Sekyere Afram Plains | 1 |
| Sekyere Central | 2 |
| Sekyere East | 2 |
| Sekyere Kumawu | 1 |
| Sekyere South | 4 |
| Asokore Mampong | 2 |
| Asokwa Municipal | 1 |
| Kumasi Metro | 5 |
| Kwadaso Municipal | 2 |
| Old Tafo Municipal | 2 |
| Suame Municipal | 2 |
| Oforikrom Municipal | 3 |
