## Supplementary material for "Availability and Use of Mobile Health Technology for Disease Diagnosis and Treatment Support by Health Workers in the Ashanti Region of Ghana: A Cross-sectional Survey": survey tool

**University of Kwazulu-Natal, Durban, School of Nursing and Public Health, Discipline of Public Health Medicine**

***Title: Mobile health (mHealth) technology for disease diagnosis and treatment support by health professionals in Ghana***

Thank you for accepting to participate in this study. This questionnaire has three sections:

Part I: Demographic information

Part II: Availability of mHealth for diagnostics and treatment support

Part III: Use of mHealth for diagnostics and treatment support

***Mark with an X in the appropriate box***

**PART I: DEMOGRAPHIC INFORMATION**

| Number | Question | Response |
| --- | --- | --- |
| 1. | Age | 20-30 years  31-40 years  41-50 years  51-60 years |
| 2. | Sex | Male  Female |
| 3. | Categories of health professionals | Medical Doctor  Physician Assistant  Midwife  General Nurse  Community Health Nurse  Laboratory Scientist/technician  Pharmacist/Dispensary technicians  Others (Specify) _______________ |
| 4. | Type of facility | District Hospital  Sub-district health center  Clinic  CHPS |
| 5. | Total number of healthcare professionals | _______________________ |
| 6. | How many patients do you see per week? | _______________________ |

**PART II: AVAILABILITY OF MOBILE HEALTH FOR DIAGNOSTICS AND TREATMENT SUPPORT**

| ***Section A: Available Health Infrastructure*** | | |
| --- | --- | --- |
| **Number** | **Question** | **Response** |
| 7. | Do you or your facility have mobile wireless devices to support the provision of healthcare? | Yes  No |
| 8. | Are there mobile health (mHealth) interventions available in this facility to support healthcare delivery? | Yes  No  ***If No (Go to Section D)*** |
| 9. | What are the various types of mobile health interventions available to health professionals in this facility?  *Tick all that apply* | Text message  Voice/phone calls  Mobile apps  Multimedia messaging    Video conferencing  Emergency toll-free lines  Others (Specify) ___________  ____________ |
| 10. | What are the various types of wireless devices available to health professionals for mobile health interventions in this facility?  *Tick all that apply* | Mobile phones  Smartphones  Tablets  Personal digital assistants  Handheld devices  Patient monitoring devices  Watches |
| 11. | Do you have continuous supply of power supply to support mobile health interventions in this facility? | Yes  No |
| 12. | Are there support systems available for the existence of mHealth for diagnostics and treatment support? | Yes  No  ***If Yes list them*** ___________  ___________________  ___________________  ____________________  ____________________ |
| ***Section B: Healthcare Workforce Competency*** | | |
| 13. | Do you have the requisite skills to use mHealth interventions for diagnostic purposes? | Yes  No  **If Yes list the skills ________**  **______________________**  **_______________________**  **_______________________**  **________________________** |
| 14. | Do you have the competence to use mHealth interventions to treat, monitor and manage diseases? | Yes  No |

**PART III: USE OF MOBILE HEALTH FOR DIAGNOSTICS AND TREATMENT SUPPORT**

| ***Section C: You and Your Mobile Wireless Device*** | | |
| --- | --- | --- |
| 15. | Have you ever used or currently using mHealth interventions to support healthcare delivery? | Yes  No |
| 16. | What type of disease(s) has mHealth been used or is currently being used for in this facility?  *Tick all that apply.* | HIV  TB  Hypertension  Diabetes  Stroke  Cancer  Chronic Respiratory disease  Malaria  Diarrhoea  Others (Specify) ____________    ____________ |
| 17. | Have you ever used smart mobile wireless device to:  *Tick all that apply.* | Find health or medical information  Disease diagnosis  Treat and manage disease conditions  Treat and monitor patients’ health conditions |
| 18. | How often do you use mobile wireless device for diagnostic purposes? | Once a month  2 or 3 times a month  1 to 6 times a week  Once a day or more |
| 19. | How often do you use mobile wireless device for treatment, monitoring and management of diseases? | Once a month  2 or 3 times a month  1 to 6 times a week  Once a day or more |
| ***Section D: Usefulness of mobile health interventions*** | | |
| 20. | Do health professionals use mHealth to monitor patients’ disease conditions? | Yes  No |
| 21. | Do health professionals use mHealth to manage non-communicable diseases like diabetes, hypertension etc? | Yes  No |
| 22. | Do health professionals use mHealth to manage communicable diseases such as HIV, TB etc? | Yes  No |
| 23. | Do health professionals use mHealth as reminders to improve their treatment adherence procedures? | Yes  No |
| 24. | Do health professionals use mHealth as reminders to promote patients’ medication adherence? | Yes  No |
| 25. | Do health professionals use mHealth to remind patients to honour their clinic appointments? | Yes  No |
| 26. | Do health professionals use mHealth to remind patients to collect their ART and other drugs on time? | Yes  No |
| 27. | Do health professionals use mHealth for follow-ups to promote treatment compliance? | Yes  No |
| 28. | Do health professionals use mHealth to support patients test result notifications? | Yes  No |
| 29. | Does the use of mHealth improve the treatment and management of disease conditions? | Yes  No |
| 30. | Do health professionals use mHealth for making accurate diagnostic decisions? | Yes  No |
| 31. | Does the use of mHealth intervention increase the effectiveness of treatment and management of diseases? | Yes  No |
| ***Section E: Ease of Use*** | | |
| 32. | Is it easy to use mobile health interventions to support disease diagnosis? | Yes  No |
| 33. | Is it easy to use mobile health interventions to support the treatment of patients’ disease conditions? | Yes  No |
| 34. | Is it flexible to interact with mobile health devices for disease diagnosis and treatment support? | Yes  No |
| 35. | Is it frustrating to interact with mobile health devices for disease diagnosis and treatment support? | Yes  No |
| 36. | Is it easy for me to become skilful in using mHealth for disease diagnosis and treatment support? | Yes  No |
| 37. | Is it easy for me to learn how to use mobile health devices for diagnosis and treatment support would be easy for me? | Yes  No |
| ***Section F: User Satisfaction*** | | |
| 38. | Do you feel comfortable in using mobile health for disease diagnosis and treatment procedures? | Yes  No |
| 39. | Are you confident in using mobile health for disease diagnosis and treatment procedures? | Yes  No |
| 40. | Are you completely satisfied in using mobile health for disease diagnosis and treatment procedures? | Yes  No |
| 41. | Do you believe that using mHealth for disease diagnosis and treatment support will increase the quality of healthcare delivery? | Yes  No |
| ***Section G: Behavioural Intention to Use*** | | |
| 42. | Would you use mHealth for the treatment and management of patients’ disease conditions? | Yes  No |
| 43. | Would you as a health professional, always use mHealth for disease diagnosis and treatment support? | Yes  No |
| 44. | If you have access to mHealth, do you intend to use it for disease diagnosis and treatment support? | Yes  No |

**Thank you for your cooperation**
